# Impact of vaccination and non-pharmaceutical interventions on SARS-CoV-2 dynamics in Switzerland

**DOI:** 10.1101/2021.04.14.21255503

**Authors:** Andrew J. Shattock, Epke A. Le Rutte, Robert P. Dünner, Swapnoleena Sen, Sherrie L. Kelly, Nakul Chitnis, Melissa A. Penny

**Author notes:** Contributed equally (Andrew J. Shattock, Epke A. Le Rutte, Robert P. Dünner).

## Abstract

As vaccination coverage against SARS-CoV-2 increases amidst the emergence and spread of more infectious and potentially more deadly viral variants, decisions on timing and extent of relaxing effective, but unsustainable, non-pharmaceutical interventions (NPIs) need to be made. An individual-based transmission model of SARS-CoV-2 dynamics, OpenCOVID, was developed to compare the impact of various vaccination and NPI strategies on the COVID-19 epidemic in Switzerland. We estimate that any relaxation of NPIs in March 2021 will lead to increasing cases, hospitalisations, and deaths resulting in a ‘third wave’ in spring and into summer 2021. However, we find a cautious phased relaxation can substantially reduce population-level morbidity and mortality. We find that faster vaccination campaign can offset the size of such a wave, allowing more flexibility for NPI to be relaxed sooner. Our sensitivity analysis revealed that model results are particularly sensitive to the infectiousness of variant B.1.1.7.

## Introduction

The COVID-19 pandemic has caused a public health and economic crisis worldwide. With rollout of COVID-19 vaccines following approval in December 2020 and in early 2021, there are many questions about when and how to relax non-pharmaceutical interventions (NPIs) while continuing to best protect the population. SARS-CoV-2 began emerging in Switzerland over 12-months ago, and by the end of January 2021 approximately 8,725 deaths were reported^1^. In response to the first steep increase in cases (the ‘first wave’) in the spring of 2020, a variety of NPIs were introduced, such as physical distancing, contact tracing, isolation of contacts, quarantining of confirmed cases, and closure or limited openings of shops and schools, with facemasks being introduced later. As a result, COVID-19 case numbers, ICU admissions, and deaths decreased prior to the summer of 2020, which led to a relaxation of some NPIs. In October 2020, Switzerland experienced a major second wave of infections, as did other countries in Europe, and NPIs were again strengthened^2^. The social and economic consequences of certain NPIs make them unsustainable in the long-term, while emergence of more transmissible SARS-CoV-2 variants are presenting new challenges. However, rollout of safe and efficacious vaccines will likely contribute to a substantial reduction in pandemic burden and thus raises the possibility of relaxing current NPIs while still potentially protecting the health and wealth of the population.

Mathematical transmission models can provide insights to support decision-making around different epidemic control strategies and public health objectives^3–5^. We present an individual-based model, ‘OpenCOVID’, which captures SARS-CoV-2 transmission dynamics using an age-structured population network that includes risk-groups (e.g., elderly and healthcare workers) and seasonal patterns. Figure 1 shows the disease and care states represented in OpenCOVID. The model was calibrated to Swiss epidemiological data – including data on variants of concern (VOC) – from 18 February 2020 to 5 March 2021. The model is described briefly in this manuscript, with additional details in the Supplementary Information.

**Fig. 1:**
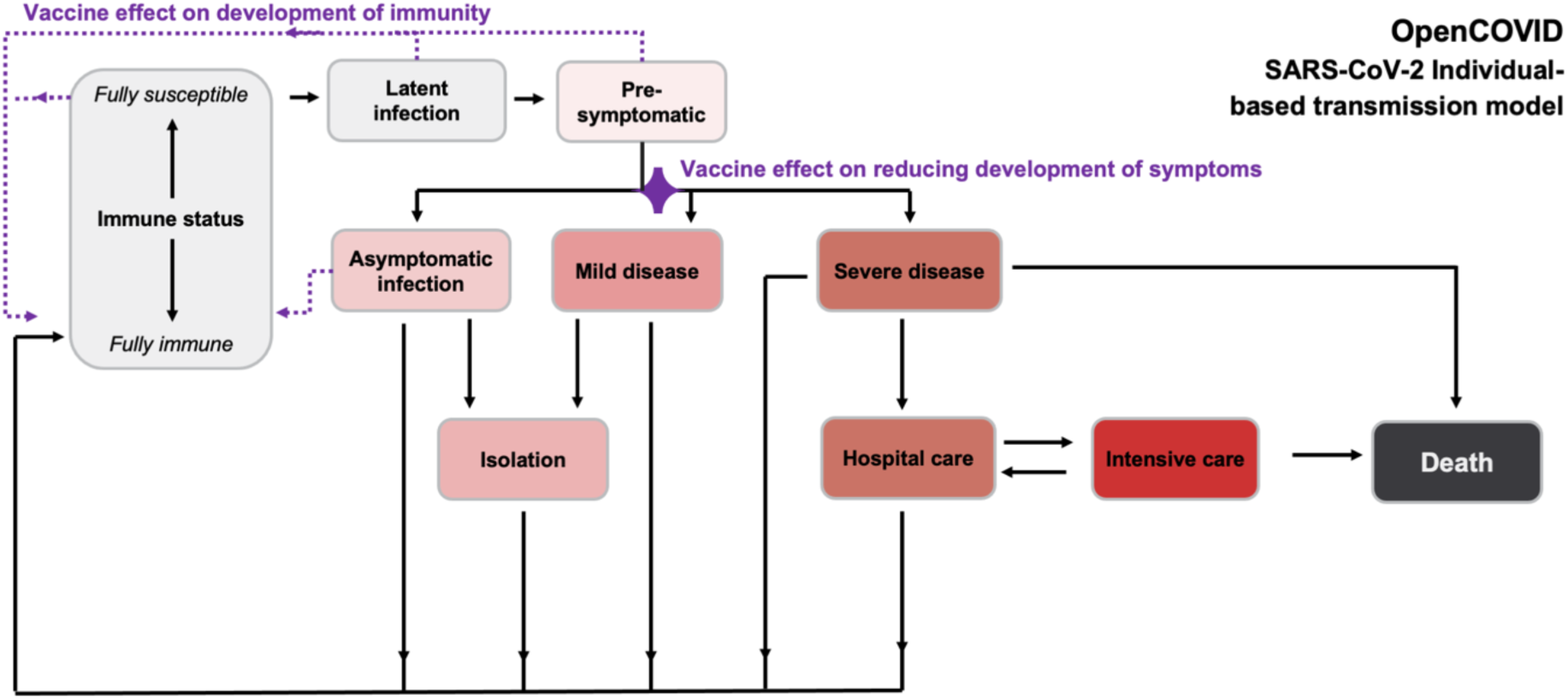
Schematic of OpenCOVID model structure. Capturing potential states of individuals in the model, including fully susceptible or immune status (exposure), latent infection, pre-symptomatic state for which vaccination may affect immune development (indicated by dotted purple lines). Some remain asymptomatic, while disease progression may occur (mild, severe) for which vaccination could reduce symptom development. Isolation or care (hospital care, intensive care) may be required. Recovery (from symptomatic infection, mild or severe disease) or death results. Increasingly darker shading (grey, pink, red, dark grey) indicates increasing severity. Full model details are provided in the Methods and Supplement.

We modelled multiple vaccine rollout scenarios with several phased NPI relaxation strategies and examined the interplay and potential impact on the epidemic in Switzerland. The impact of NPIs was modelled as a reduction in effective contacts scaled using the Oxford Containment and Health Index^6^. Vaccine rollout was modelled according to the strategy defined by the Swiss Federal Office of Public Health (FOPH). This analysis was conducted in early March 2021 prior to a Federal decision for potential relaxation of prevention measures planned to begin 22 March. The model and scenario analyses were done at the national level, and therefore do not capture the substantial heterogeneity within or between Swiss cantons. We explored when and by how much NPIs could be relaxed alongside two speeds of vaccine rollout to prevent or limit a potential ‘third wave’ surge of confirmed cases, hospitalisations, ICU admissions, and deaths. Since factors other than control measures can strongly influence the course of the epidemic, using a sensitivity analysis we examined the potential impact of 1) varying properties of VOCs, 2) varying vaccine efficacies, and 3) varying levels of vaccine uptake in the population. Potential future changes or developments in the clinical care and general population health were not considered here; nor were potential changes in mass testing or in rates of effective testing, tracing, isolating, and quarantining, or the introduction of new, as of now, interventions.

## Results

OpenCOVID was calibrated to national-level epidemiological data up to 5 March 2021 and accurately captured historical trends of confirmed cases, hospital and ICU cases, and deaths. See Supplementary Figures S1 and S2, also Figure 2. By the end of February 2021, we estimated that 18%–26% of the Swiss population had been exposed to SARS-Cov-2, approximately aligned with regional seroprevalence surveys^7, 8^. It was assumed that variant B.1.1.7 has 60% more transmissibility than D614G^9^, with which OpenCOVID accurately captured available genomic surveillance data in Switzerland^10–13^ (Supplementary Figure S3). For control measures in place from January to February 2021, this increased transmissibility translates to a relative transmission advantage of 1.3–1.4 over this period.

**Fig. 2:**
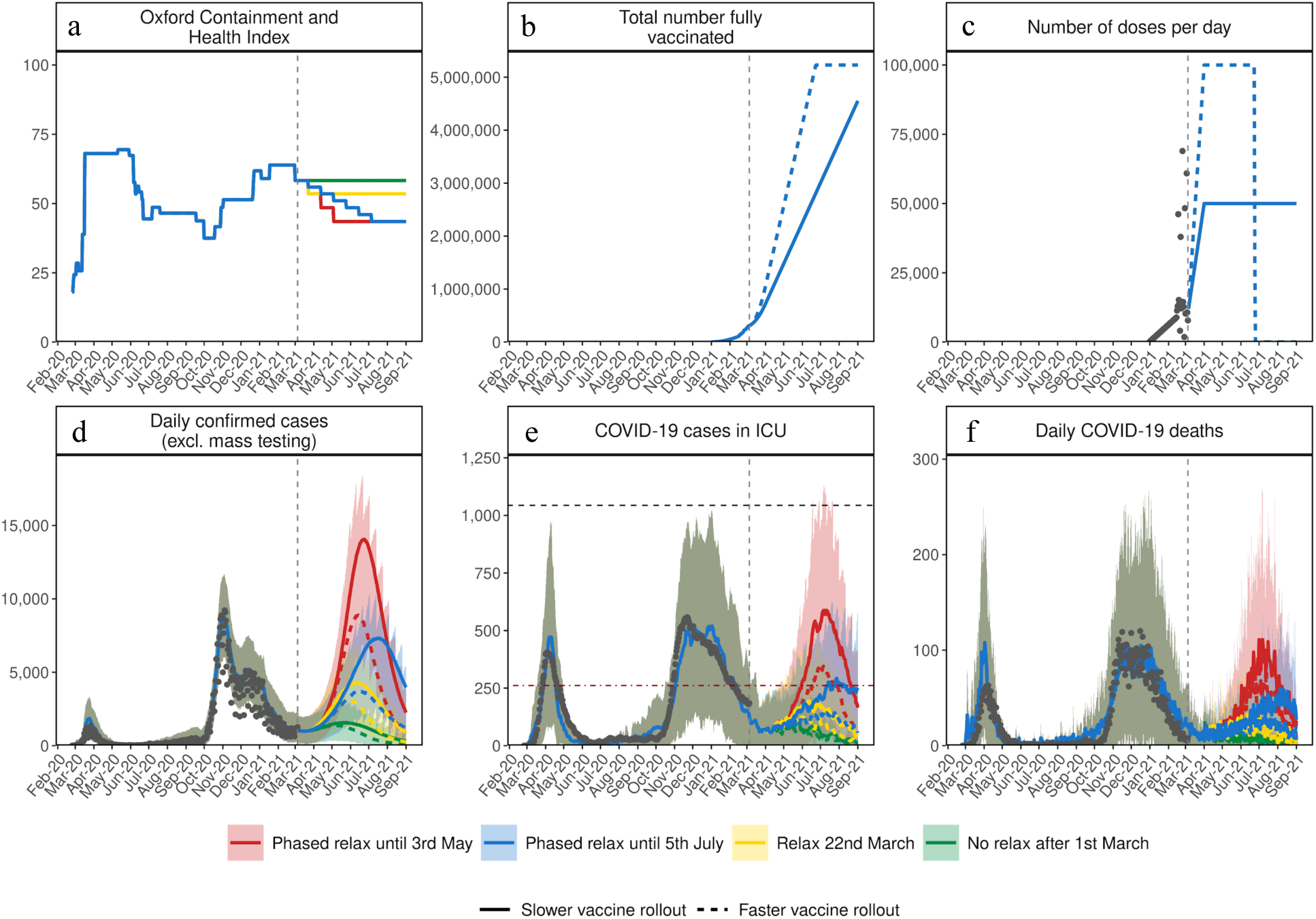
Estimated impact of vaccination and NPI relaxation scenarios on the SARS-CoV-2 epidemic in Switzerland. (**a**) Oxford Containment and Health Index: A measure for the severity of NPIs from 24 February 2020 to 21 March 2021 and projections for four relaxation scenarios thereafter. (**b**) Total number fully vaccinated: Cumulative amount of fully vaccinated persons (assuming two doses). (**c**) Number of doses per day: Number of vaccine doses administered per day. (**d**) Daily confirmed COVID-19 cases: Model estimates of the number of confirmed COVID-19 cases per day (not accounting for future testing changes including mass testing). (**e**) COVID-19 cases in ICU: Model estimates of COVID-19 patients in ICU. (**f**) Daily COVID-19 deaths: Model estimates of daily COVID-19-related deaths. In all panels, dark grey dots show data to date. Coloured lines show simulation results of different relaxation scenarios and two vaccine rollout scenarios. Vaccination scenarios: Solid lines correspond to a vaccination scenario assuming 50,000 vaccines are administered per day, while the dashed lines correspond to a faster vaccination scenario of 100,000 vaccines per day. NPI scenarios (details Table 1): Red represents an NPI relaxation scenario with relaxation steps on 22 March, 12 of April, and 3 of May. Blue represents a slower NPI relaxation scenario compared with the red scenario, with smaller relaxations, three-weekly steps from 22 March to 5 July. Yellow represents an NPI relaxation scenario with relaxation only on 22 March and no further NPI relaxations. Green represents a strategy with no further NPI relaxation after 1 March (an unrealistic scenario of no relaxations through to the end September 2021, provided as a reference only). The vertical dashed lines represent the date at time of analysis. The horizontal black dashed line in the “COVID-19 cases in ICU” panel depicts the estimated maximum national capacity of ICU beds; the horizontal red dashed line 25% ICU capacity. Predictions of confirmed cases, ICU capacity, and mortality are reported as mean estimates with 95% prediction intervals.

**Table 1:** Summary of model scenarios from Figures 2 and 3. Oxford Containment and Health Index (OCHI) levels are detailed in the methods.

| Scenario | NPI relaxation speed | NPI OCHI level on 22 March | NPI OCHI level on 12 April | NPI OCHI level on 3 May | NPI OCHI level on 5 July | Vaccination speed | VOC transmission | Vaccine assumptions |
| --- | --- | --- | --- | --- | --- | --- | --- | --- |
| 1A) RED | NPI relaxation steps on 22 March, 12 April, and 3 May 2021 | 53.5<br>Similar to levels in early June 2020 | 48.5<br>Similar to levels in early July 2020 | 43.5<br>Similar to levels at the end September 2020 | 43.5 | 50,000 per day (solid line) or 100,000 per day (dashed line) | Baseline – 60% increased transmission | Baseline - 80% transmission blocking |
| 1B) BLUE | Small NPI relaxations every 3-weeks between 22 March and 5 July 2021 | 55.9<br>Similar to levels in early June 2020 | 53.5<br>Similar to levels in early June 2020 | 51.0<br>Similar to levels in November and December 2020 | 43.5<br>Similar to levels at the end September 2020 | 50,000 per day (solid line) or 100,000 per day (dashed line) | Baseline – 60% increased transmission | Baseline – 80% transmission blocking |
| 1C) YELLOW W | NPI relaxation step on 22 March 2021 with no further NPI relaxations (in accordance with relaxations communicated on 17 February) | 55.9<br>Similar to levels in early June 2020 | 55.9 | 55.9 | 55.9 | 50,000 per day (solid line) or 100,000 per day (dashed line) | Baseline – 60% increased transmission | Baseline – 80% transmission blocking |
| 1D)<br>GREEN | No further relaxation after 1 March 2021 | 58.3<br>Since 1 March 2021 | 58.3<br>Since 1 March 2021 | 58.3<br>Since 1 March 2021 | 58.3<br>Since 1 March 2021 | 50,000 per day (solid line) or 100,000 per day (dashed line) | Baseline – 60% increased transmission | Baseline – 80% transmission blocking |

Findings from our scenario analyses suggest that even if NPIs in place in early March 2021 are maintained until September 2021 (an unrealistic scenario), daily cases, hospitalisations, and deaths will increase (Figures 2 and 3), resulting in a third wave. Any relaxation during the month of April is also predicted to lead to increases in all three indicators (Figures 4 and 5). This is due to 1) increased rates of human contact following relaxation of NPIs, 2) the majority of the population still being immunologically naïve (i.e., no prior infection or vaccination, Supplementary Figure S10), and 3) the increasing incidence of VOC with higher transmissibility. Since these findings may be sensitive to uncertainties in VOC properties these factors were further explored via sensitivity analysis. Scenario details and assumptions are provided in Tables 1 and 2.

**Fig. 3:**
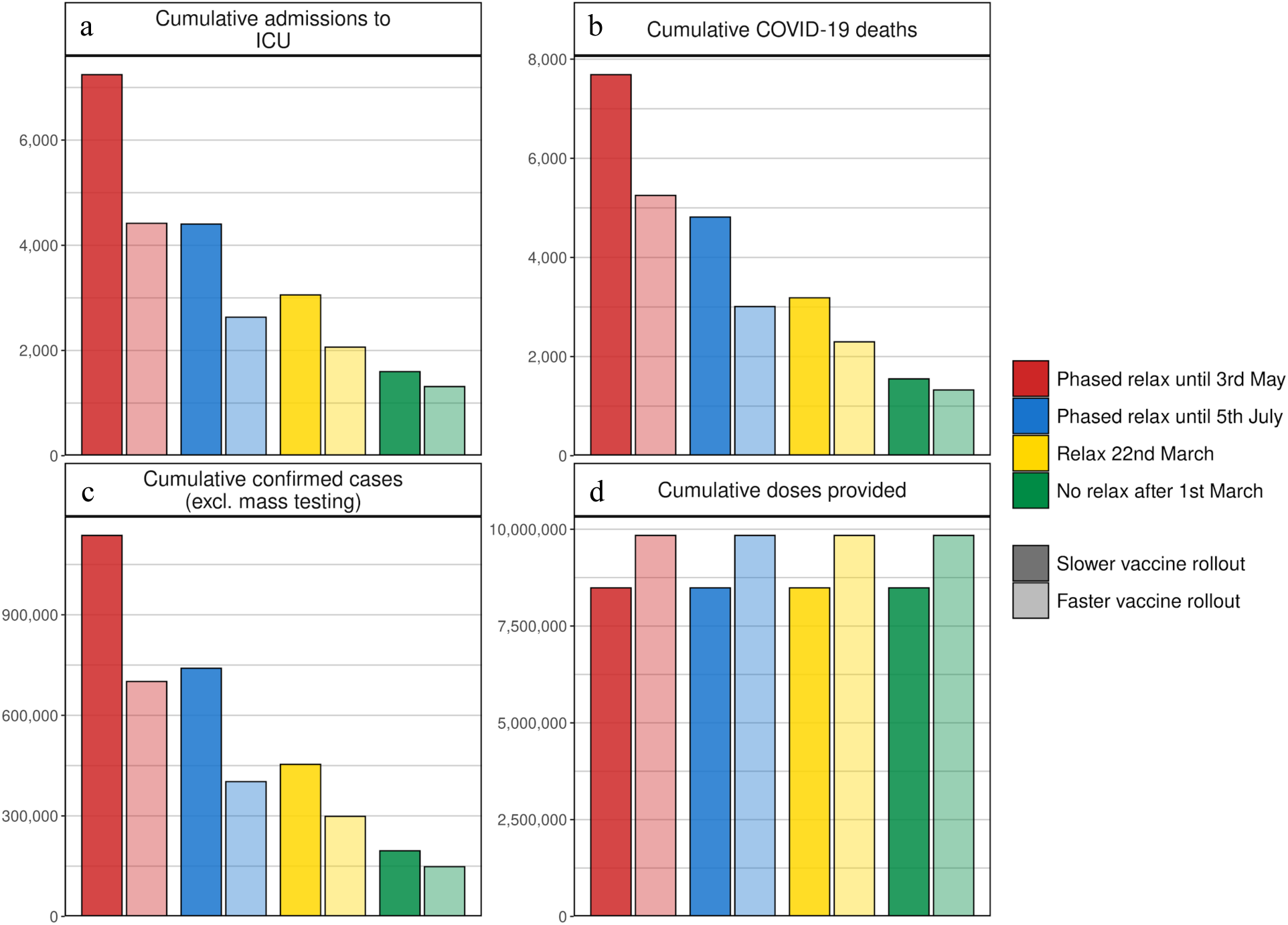
Cumulative estimates of confirmed cases, ICU admissions, COVID-19 deaths, and vaccine doses provided from 6 March to 5 September 2021. (**a**) Cumulative admissions to ICU, (**b**) cumulative COVID-19 deaths, (**c**) cumulative confirmed COVID-19 cases (excl. mass testing), (**d**) cumulative vaccine doses provided. Bar colours indicate NPI relaxation and vaccination scenarios as per Figure 2, with darker shaded bars correspond to vaccination scenarios with 50,000 vaccines administered per day and lighter shaded bars to faster vaccination scenarios with 100,000 vaccines administered per day.

**Fig. 4:**
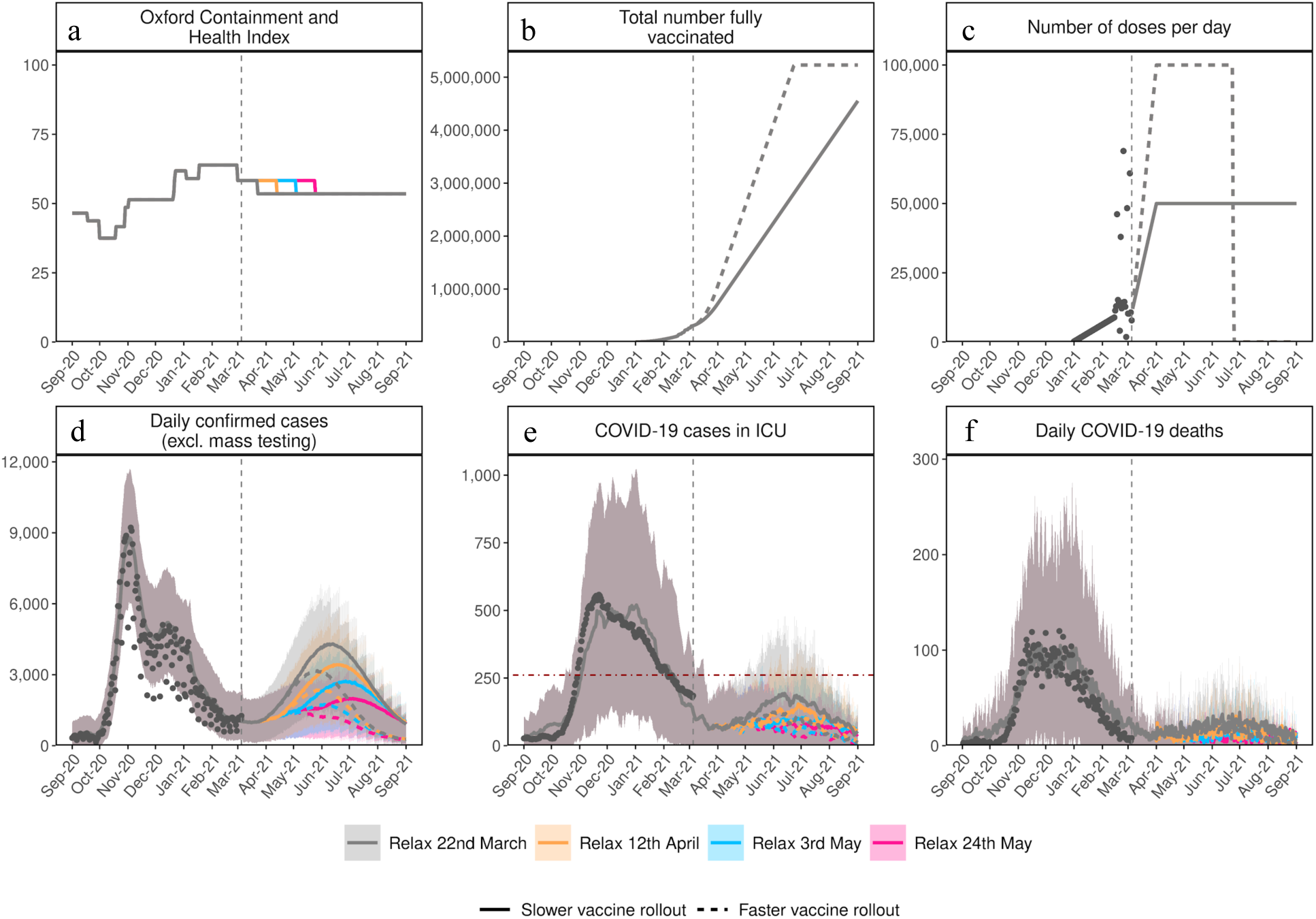
Comparison of the impact of vaccination and NPI relaxation scenarios on SARS-CoV-2 dynamics in Switzerland over time for delayed single-step relaxation. (**a**) Oxford Containment and Health Index: A measure for the severity of NPI measures from 24 February 2020 until 21 March 2021 and for four exemplar relaxation scenarios with one step relaxation. NPI scenarios: Grey represents an NPI relaxation scenario with relaxation only on 22 March with no further NPI relaxation (also detailed as the yellow scenario in Table 1); Orange same relaxation but implemented three- weeks later on 12 April; Blue same relaxation on 3 May, and Pink same relaxation 24 May. (**b**) Total number fully vaccinated: Cumulative amount of fully vaccinated persons (assuming two doses). (**c**) Number of doses per day: Number of vaccine doses administered per day. (**d**) Daily confirmed cases: Model estimates of the number of confirmed COVID-19 cases per day (not accounting for future testing changes including mass testing). (**e**) COVID-19 cases in ICU: Model estimates of COVID-19 patients in ICU. (**f**) Daily COVID-19 deaths: Model estimates of daily COVID-19-related deaths. In all panels dark grey dots show data to date. The coloured lines show simulation results for the different relaxation scenarios and the two vaccine rollout scenarios. Vaccination scenarios: Solid lines correspond to a vaccination scenario assuming 50,000 vaccines are administered per day, while dashed lines correspond to a faster vaccination scenario of up to 100,000 vaccines per day. The vertical dashed line on all panels represents the current date at the time of the analysis. The horizontal red dashed line in the “COVID-19 cases in ICU” panel depicts the level of 25% of ICU capacity. Predictions of confirmed cases, ICU capacity, and deaths are mean estimates with 95% prediction intervals.

**Fig. 5:**
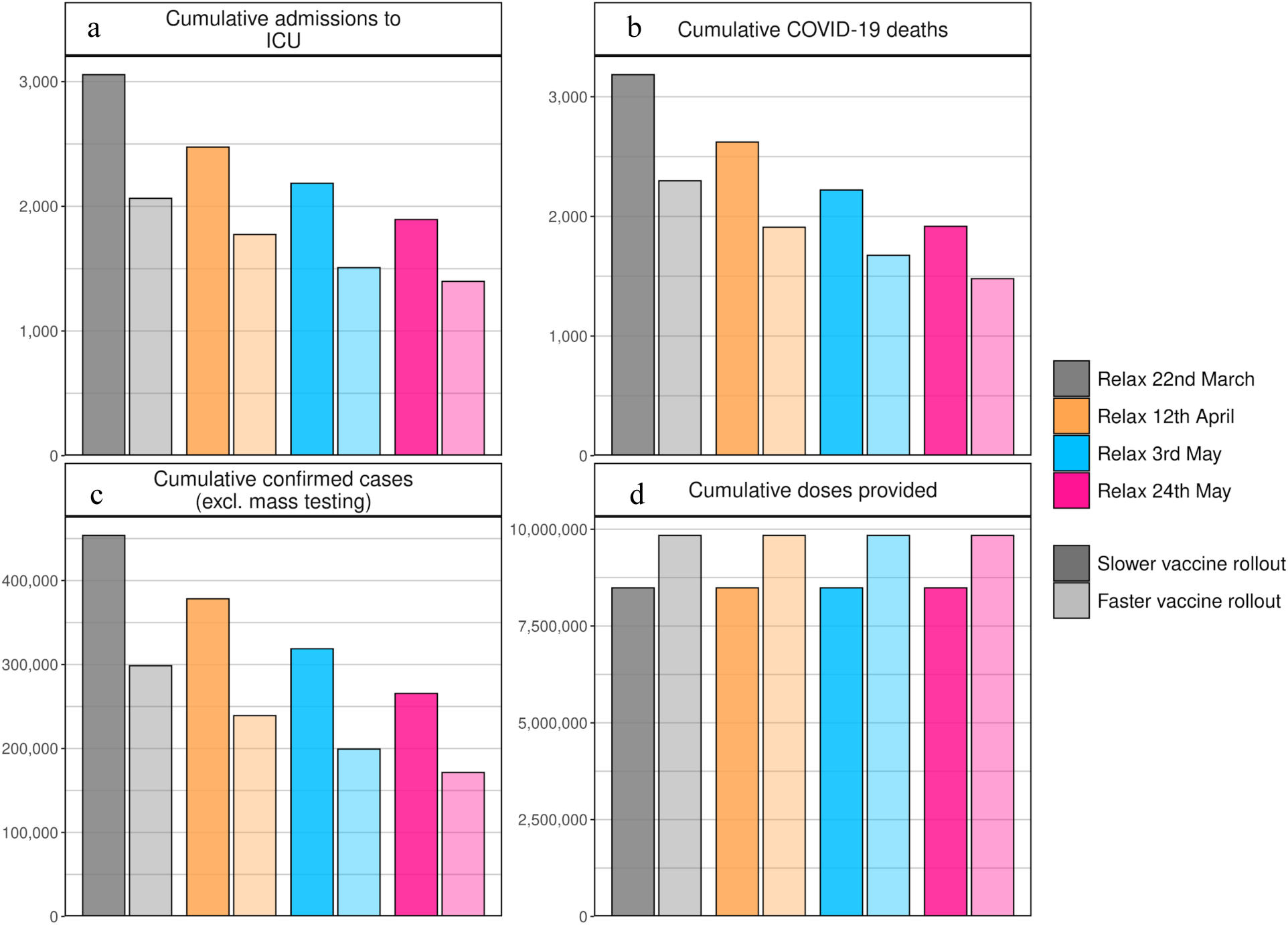
Cumulative estimates of the number of confirmed cases, ICU admissions, COVID-19 deaths, and vaccine doses provided between 6 March and 1 September 2021. (**a**) Cumulative admissions to ICU, (**b**) cumulative COVID-19 deaths, (**c**) cumulative confirmed COVID-19 cases (excl. mass testing), (**d**) cumulative vaccine doses provided. Bar colours indicate NPI relaxation and vaccination scenarios as per Figure 4 with darker shaded bars correspond to the slower vaccination scenario with 50,000 vaccinations per day and lighter shaded bars to the faster vaccination with100,000 vaccinations per day.

**Table 2:**
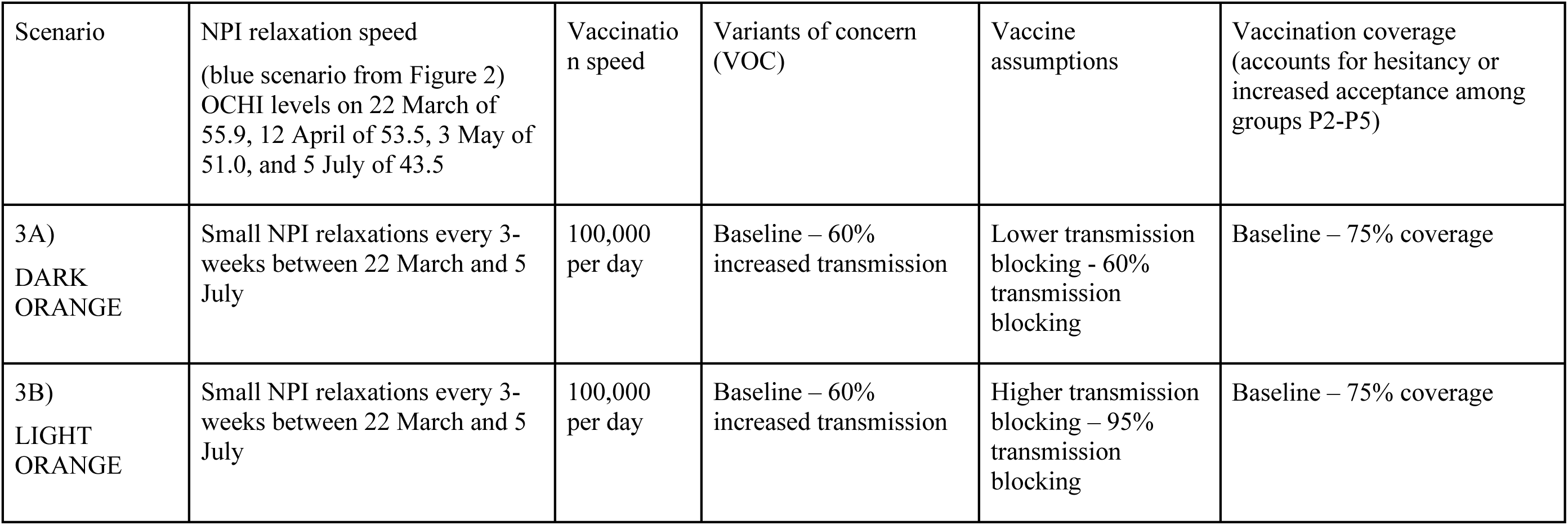
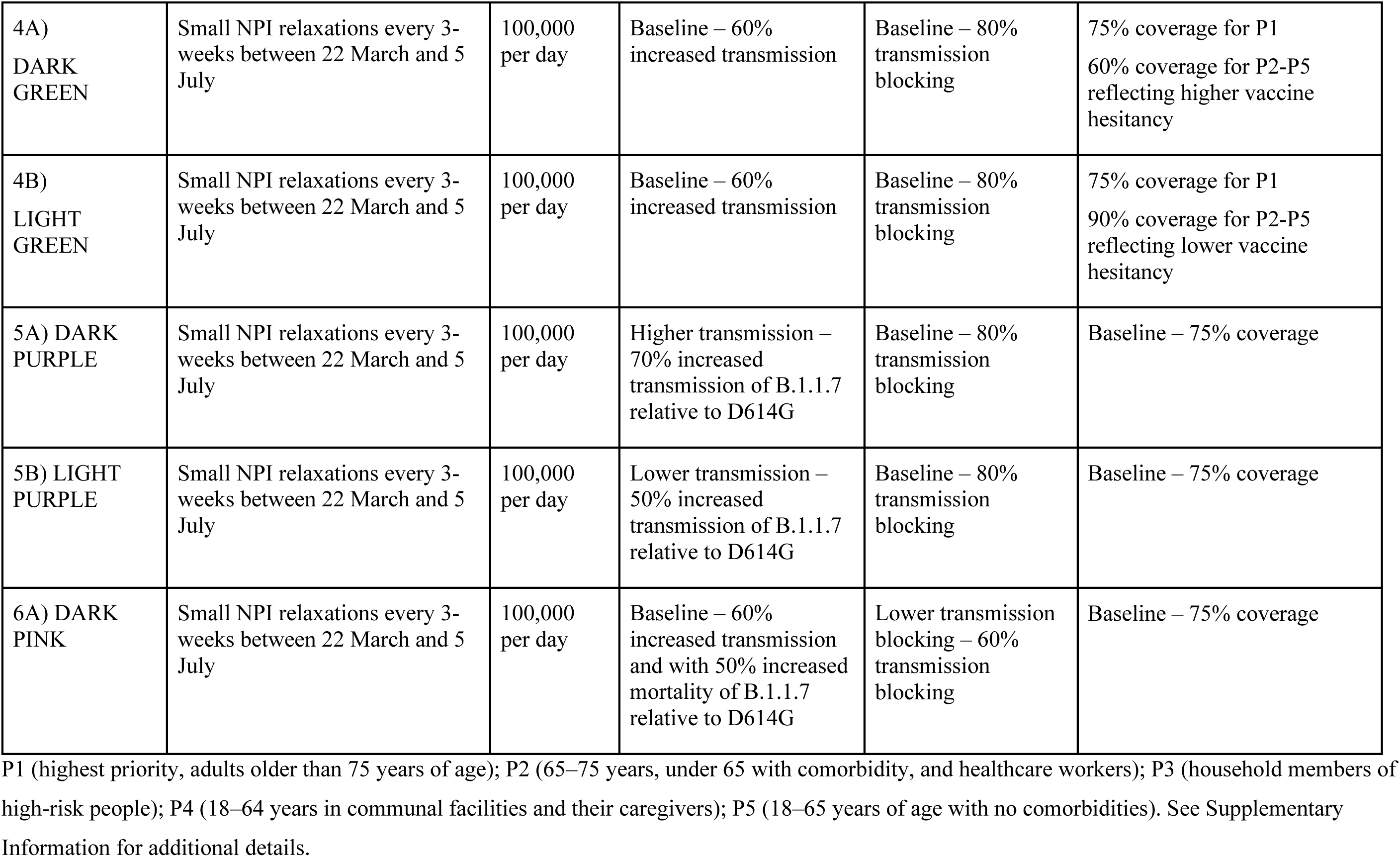
Summary of model scenarios from Figures 6 and 7

The size of the resulting third wave is strongly dependent on timing and amount of measure relaxation. Faster and stronger relaxations lead to larger third waves, while slower relaxations resulted in a smaller, but also a delayed third wave peak (Figure 2). If relaxation is particularly strong, either in large steps or via quick successive relaxations (e.g., the ‘red’ scenario), we estimate that ICU occupancy will increase above 25% capacity, a key indicator for decision makers^14^. The size of a potential third wave however can be substantially reduced with faster vaccination by increasing the number of individuals vaccinated per day. We found that increasing vaccination rates from 50,000 to 100,000 doses used per day (0.6% and 1.2% of the Swiss population, respectively) results in a halved and slightly earlier third wave peak. Furthermore, for the more gradual phased relaxation scenario, this increased vaccination rate results in a substantial reduction in ICU occupancy and deaths until September 2021.

The impact of delaying NPI relaxation was explored by comparing a relaxation on 22 March with the same relaxation delayed by 3-, 6-, or 9-weeks (Figure 4). Simulations indicated that delaying relaxation could lead to fewer cases, less morbidity, and less mortality. However, similar gains could also be achieved through faster vaccination, facilitating a more flexible epidemic exit strategy.

For many scenarios, we observe a peak in cases and ICU occupancy in the summer of 2021 followed by a decrease. The initial rise in cases occurs through infections of people who were not previously infected or vaccinated rather than from re-infection of previously infected individuals or due to imperfect vaccine efficacy (Figure S5). Scenarios involving quicker NPI relaxation led to more person-to-person contact, increased transmission, and faster population immunity, which along with vaccination, build-up incrementally until the summer of 2021 (Figures 2 and 3). This also results in a large wave of infections, high ICU occupancy, and deaths in spring and into summer 2021 (Figures 2 and 3). The peak in cases occurs when there are a sufficient number of people with vaccine-induced or natural immunity due to infection, such that while NPI measures are in place transmission is largely decreased. The subsequent decay in new daily cases occurs because fewer individuals are susceptible, and along with NPI measures remaining in place, results in an effective reproduction number of less than one. When projections increase towards Switzerland’s maximum ICU capacity or to death rates as high as observed in previous waves, predicted trajectories become highly unlikely since additional measures would likely be implemented.

Assumptions around increased transmission of and mortality from new VOC, vaccine properties, and vaccine hesitancy were further explored. The biggest driver of impact from vaccination and NPI relaxation on mortality was the level of increased transmission of VOC (Figures 6 and 7). If new variants are more transmissible than assumptions explored here (namely greater than 70%), then there is a considerable risk of overwhelming the health system. Furthermore, if new variants have a 50% increased mortality compared with D614G^15, 16^, then 41% to 44% more deaths may occur from March 2021 to September 2021, depending on the NPI relaxation strategy and the speed of vaccination. Even with faster vaccination, if partial onward transmission from individuals who are vaccinated but then become infected occurs (i.e., less than 65% transmission blocking is assumed), an even slower NPI relaxation is needed to avoid subsequent re-strengthening of measures in the future. Regardless of vaccination speed, with higher vaccine acceptability among groups P2-P5 (90%), fewer cases are predicted during the third peak of the epidemic, compared with 60% lower acceptability (Figure 6).

**Fig. 6:**
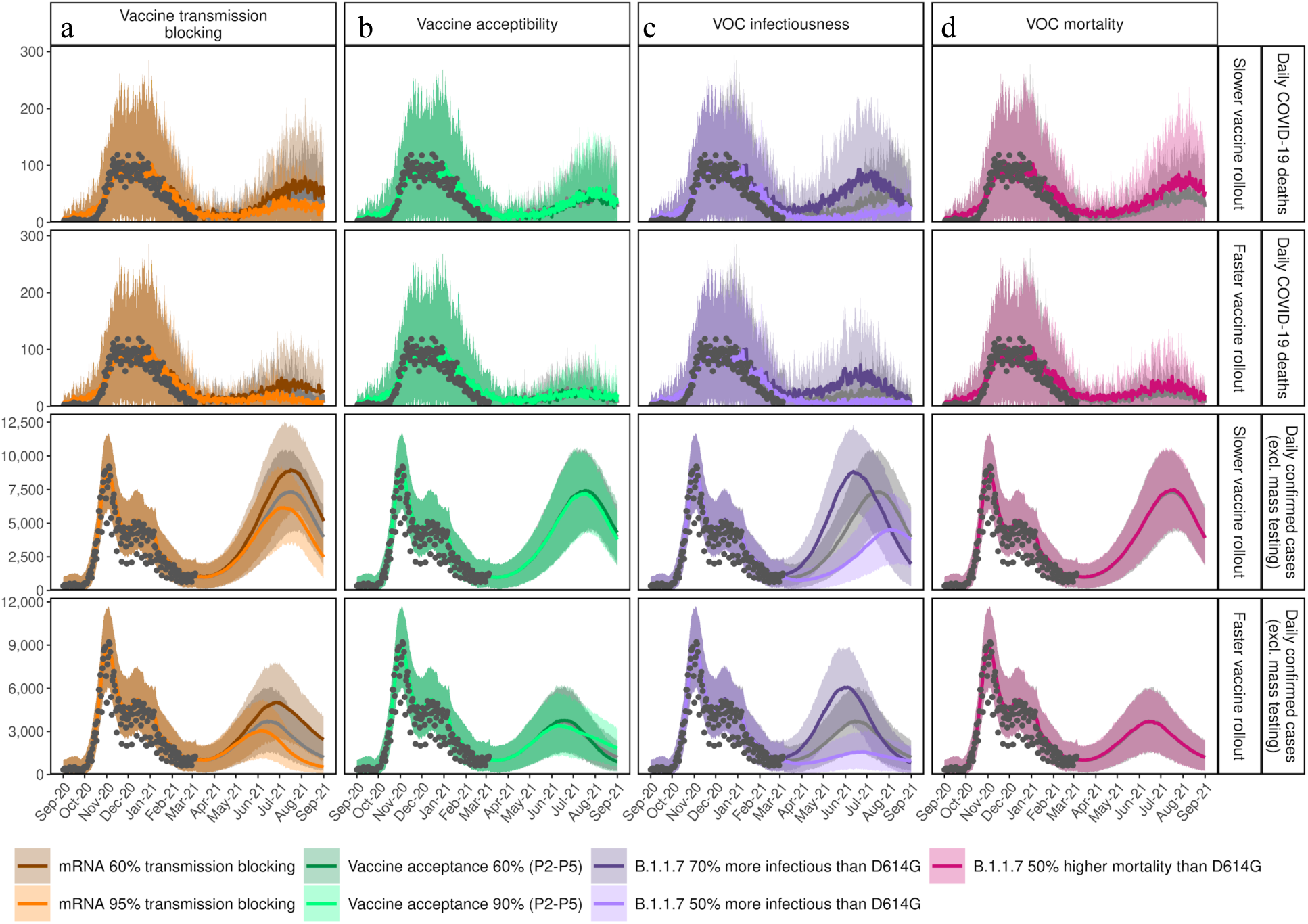
Sensitivity of predictions for daily confirmed cases and COVID-19 deaths given assumptions for vaccine transmission blocking, vaccine acceptability, infectiousness of VOC, and potential increased mortality from VOC. Each panel shows the time series of either daily confirmed COVID-19 cases (excluding testing changes) or daily COVID-19 deaths (indicated by the row labels) between 1 September 2020 and 1 September 2021 for the blue scenario with slower relaxations every three-weeks from 22 March to 5 July with 50,000 vaccines per day (rows 1 and 3) or faster vaccination with 100,000 per day (rows 2 and 4). Results of the blue scenario are represented by grey dots, and colours show the impact of (**a**) vaccine transmission blocking, influence of the assumption on the transmission blocking property of the vaccine. Dark orange line represents 60% vaccine transmission blocking, light orange line 95% transmission blocking, and the grey line the best estimate for 80% transmission blocking used in Figures 2 and 3. The transmission blocking property of the vaccine is offset with the symptom blocking property, so that the vaccine always has an efficacy of 95% in reducing symptoms. (**b**) vaccine acceptability in terms of coverage represents assumed vaccine hesitancy, resulting in a change in vaccination coverage. Dark green line represents 60% coverage, light green line 90% coverage, and grey line the best estimate of 75% coverage as used in Figures 2 and 3. (**c)** VOC infectiousness represents assumed increased transmission for variant Dark purple line shows a 70% increased transmissibility (with transmission advantage of 1.4– 1.5), light purple line 50% increased transmissibility (transmission advantage of 1.2–1.3), and grey line the reference scenario of 60% increased transmissibility (transmission advantage of 1.3–1.4) used in Figures 2 and 3. (**d**) VOC mortality with dark pink lines representing assumed 50% increased mortality for variant B.1.1.7 compared with D614G, while the grey lines the same mortality assumptions as for variants that emerged in 2020. Predictions of confirmed cases and mortality are reported as mean estimates with 95% prediction intervals.

**Fig. 7:**
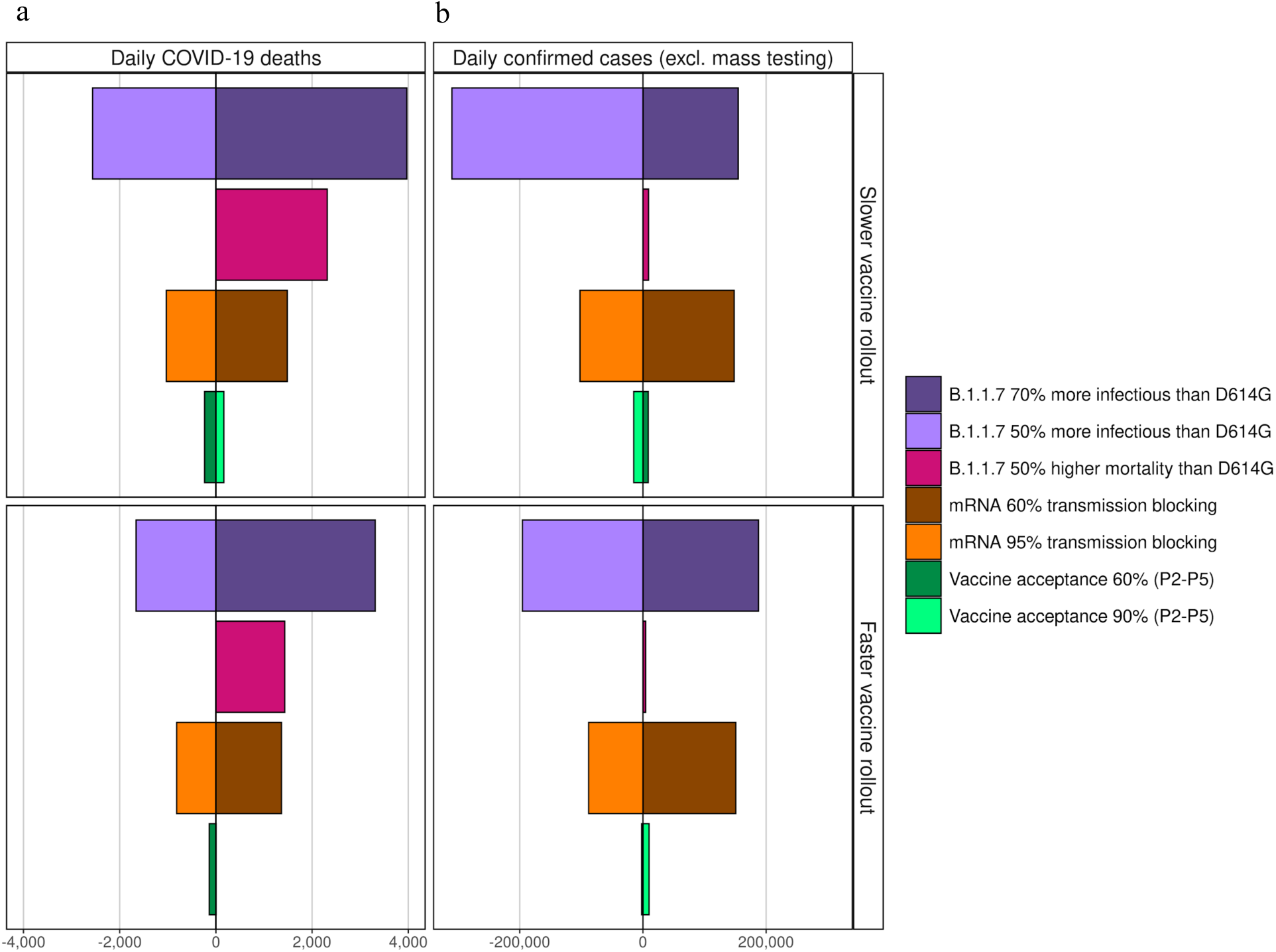
Cumulative mean impact on daily COVID-19 deaths and confirmed cases given the sensitivity analysis of Figure 6 (Table 2) on the blue reference scenario for slower relaxation until 5 July (Table 1, Figure 2). Estimated impact on (**a**) daily COVID-19 deaths and (**b**) daily confirmed COVID-19cases (excluding mass testing) from 1 September 2020 to 1 September 2021 given the different assumptions on infectiousness of VOC, potential increased mortality from VOC, and vaccine transmission blocking and vaccine acceptability as illustrated in Figure 6. The colour and shading schemes are identical to those in Figure 6. The dividing line at 0 is the reference cumulative mean estimate from Figure 3 for the dark blue scenario bar modelled with (50,000 vaccines per day) and light blue scenario bar (100,000 vaccines per day): slow phased NPI relaxation scenario of three- weekly steps from 22 March to 5 July. Influence of the assumption of the increased transmission of variant B.1.1.7: dark purple bars represent an assumed 70% increased transmissibility (corresponding to a transmission advantage of 1.4–1.5), the light purple bars, 50% increased transmissibility (corresponding to a transmission advantage of 1.2–1.3), over the reference of 60% transmissibility for D614G. Influence of the assumption of an increased mortality of 50% for variant B.1.1.7 (dark pink bars). Influence of the assumption on the transmission blocking property of the vaccine: dark orange bars represent a vaccine with 60% transmission blocking, the light orange bars represent 95% transmission blocking. The transmission blocking property of the vaccine is offset with the symptom blocking property, so the vaccine has a 95% efficacy for reducing symptoms. The influence of the assumption for vaccine hesitancy, resulting in a change of vaccination coverage: the dark green line shows a coverage of 60% (acceptance), the light green line a coverage of 90% (compared with the reference 75% coverage). For all bars, the negative values on the left indicate fewer cases or deaths are predicted compared with the reference scenario, for positive values on the right, more deaths or cases are predicted.

## Discussion

The OpenCOVID model has proven to be useful for comparing future scenarios for a range of NPI relaxation strategies, speed of vaccination rollout strategies and vaccination efficacies, as well as for examining the impact new variants on transmission dynamics. Even accounting for the uncertainty around the relative transmission advantage of new variants, all scenarios indicated that faster vaccination and more cautious, phased, NPI relaxation leads to more optimistic outcomes of COVID-19 incidence and mortality. This in turn leads to a reduced risk of surpassing 25% ICU capacity with COVID-19 patients and hence to a reduced risk of needing to re-strengthen measures in spring or summer 2021. Such a level of ICU occupancy from COVID-19 patients has the potential to fully exhaust capacity when ICU occupancy from other ailments is considered^14, 17^.

We found that significant delays exist between changing NPI measures and the measurable effect on case numbers, hospitalisations, and ICU admissions. Specifically, the consequence of a relaxation is not seen on ICU admissions until at least four- to six-weeks later. Given these delays, making decisions to relax NPI measures made more frequently than every four- to six- weeks^18^ runs the risk of causing undesirable knock-on effects, including mounting pressure on ICUs and a later need to re- strengthen measures. Furthermore, if NPIs are relaxed too frequently or too strongly before a maximum ICU capacity trigger is reached, a larger peak in ICU occupancy will occur and a stronger reactive strengthening of NPIs may be required. Decisions on trigger points for changes in NPI measures must take this delay into account to effectively control ICU occupancy.

In the coming months, vaccination alone will not be sufficient to control mortality from COVID-19. Crucially, given the increased transmissibility of the new variants, even without any further NPI relaxation after 1 March 2021, a third wave is expected, and, at the time of writing, cases were already increasing across Europe. Relaxation of NPIs is likely to lead to another wave of infection, however higher and quicker vaccine uptake, alongside a more gradual relaxation, will minimise this wave and associated mortality. Our phased relaxation scenarios indicate that if NPIs are relaxed too soon or too fast, the resulting third wave has the potential to overload the Swiss health system (e.g., red scenarios in Figure 2) and could thus prompt the need to reinforce more restrictive NPIs.

Our results suggest that to safeguard against a significant third wave and not to overwhelm the health system, rigorous monitoring of vaccination uptake and the emergence of new viral variants is required, which can inform careful decision making surrounding the timing and strength of NPI relaxations. While vaccination is ongoing, it is critical to continuously assess the impact of each NPI relaxation over a sufficient length of time before committing to additional relaxations. Effective communication with the public will be key to ensure the successful uptake of vaccines and to ensure that NPI measures, including hand hygiene, physical distancing, and facemasks, are followed to protect unvaccinated individuals and those who cannot be vaccinated. It is the combination of both vaccine uptake and ongoing NPIs that dictates the size of any third wave and timing of its peak. Even once effective vaccination campaigns end, facemasks and some level of physical distancing will likely still be required^19^, and for this reason we modelled the lowest level of NPIs to be roughly equivalent to measures that were in place in Switzerland in September 2020.

Although evidence beyond clinical studies is being accrued on vaccine efficacy (e.g., in Israel and the UK), our results are still dependent on assumptions for vaccine efficacy, uptake levels, and speed of vaccination. Over the simulated time span, we also assumed no loss of natural or vaccine-induced immunity. It will be crucial to monitor real vaccine uptake and coverage rates, as well as vaccine efficacy on new variants, and waning immunity over the coming 6- to 12-months. We modelled two vaccination strategies, a faster rollout with 100,000 doses per day and a slower rollout with 50,000 doses per day; however, the feasibility and capacity to administer these daily amounts of vaccine doses was not explored. Moreover, the impact of increasing vaccination coverage above 75% in the key risk group (adults older than 75 years of age, P1) was not explored, however, any increase in this will likely result in reduced mortality associated with the third wave. Increased vaccine hesitancy in less vulnerable groups is not expected to lead to increased mortality in the short-term. Vaccinating people under 18 years of age (since at the time of writing vaccine safety in children had yet to be tested) and delaying of second vaccine doses in order to increase coverage of first doses, or because of dosing shortages, was not explored.

We only considered variants of concern identified in Swiss genomic surveillance data ^10^; B.1.1.7, the most prevalent VOC in Switzerland as of March 2021, and B.1.351. We did not consider P.1, for which vaccines may be less efficacious^20, 21^. As more information about VOC becomes available, additional model simulations will be required to further examine the effects of new variants on the epidemic. We did not consider potential changes in behaviour among people once they are vaccinated.

We used the Oxford Containment and Health Index (OCHI) to measure the strength of NPIs. This single-value index integrates 13 different SARS-CoV-2 protection measures. Equal levels of OCHI can be reached by different combinations of NPIs. This means that the scenarios modelled are not specific to a certain defined relaxation scenario, but rather treat the effect of a certain total amount of relaxations as measured by the OCHI. The set of relaxation scenarios modelled in this study was chosen to allow exploration of interactions between different vaccination speeds and delays of relaxation, and do not represent explicit measures planned for Switzerland.

While we quantified short-term consequences of COVID-19 such as hospitalisations and mortality, we did not quantify long-term sequelae associated with severe and non-severe cases. While COVID- 19 lasts for two weeks on average^22^, an estimated one in ten individuals suffer with symptoms for more than 12 weeks^23^, defined as ‘long COVID’. Long COVID and disability associated with severe cases was not considered; however, the impact of these factors could be substantial as symptoms, including fatigue, anxiety, joint or muscle pain, and more^24^, could have a considerable impact on physical and mental health, and on workforce participation. In this context, our results were more optimistic since outcomes of long COVID and disability from severe cases were not considered. We also did not include economic consequences and secondary health impacts of relaxing and potentially re-strengthening measures. These must be considered as part of any policy decision, alongside additional economic and health analysis, and should be a priority for future modelling analyses.

As for any epidemic model, considerable uncertainty exists when projecting further into the future. The current pandemic will not end in September 2021; however, we ended our simulations at that time point due to increasing uncertainty of predicting beyond this period. During this projection period, it will be important to monitor the effect of waning immunity and any benefit from summertime climatic effects. We optimistically assumed that the seasonal effect of reduced transmission seen in summer 2020 will apply equally to the new variants of concern. It is important to highlight that the numbers of predicted cases are thus influenced by this phenomenon and that the opposite effect will take place in autumn and winter, also arguing for both high vaccination rates before autumn and effective monitoring beyond summer 2021. Precise transmission levels of new variants of concern are not yet known, which would directly affect the measure of the effective reproductive number for Switzerland. Likewise, the effectiveness of various COVID-19 vaccines on existing and possible new variants is not yet accurately known. Changes in testing (including mass testing), changes in NPI measures, and adherence to those measures may result in different future trends than those seen in 2020. Furthermore, communication of expected epidemic trends may lead to behavioural changes that make these trends less likely (such as the population becoming more careful and reducing contacts if cases are expected to increase, and vice versa). Such potential behavioural changes were not captured in the model. Although we have undertaken great effort to inform our model with the best available data and assumptions, this analysis should not be considered as a future prediction. Instead, our findings provide outcomes for various potential scenarios comparing the relative impact of different SARS-CoV-2 control strategies over the coming months at the national level and do not capture substantial heterogeneity within and between cantons.

Disease models allow us to explore counterfactual scenarios and to counterbalance the human tendency to underestimate exponential growth. The complex interaction between new emerging variants, NPI control measures, and vaccine rollout strategies is difficult to grasp without using a model. Models offer a snapshot of several possible futures. They cannot automatically react to policy changes, as these changes will make model assumptions unfounded. Instead, models provide a tool to compare the relative impact of decisions made now on the future course of the pandemic. With this study, we have addressed these policy-related considerations for the Swiss population from a public health point of view. However, these insights can be applied more broadly to countries or regions to forecast the impact response measures will have on SARS-CoV-2 transmission, which we hope will aid in global control of COVID-19.

## Methods

### Model

To represent the impact of vaccines and NPIs on the COVID-19 epidemic in Switzerland, we developed OpenCOVID; a stochastic, discrete-time, individual-based transmission model of SARS-CoV-2 infection and COVID-19 disease. OpenCOVID is briefly described here, with additional model details provided in Supplementary Information. Model code is open source and available from^25^.

OpenCOVID tracks characteristics of individuals such as age (in one-year age bins), risk-group (namely those with comorbidity), SARS-CoV-2 infection status, COVID-19 disease state, level of immunity, and vaccination details. If infected, an individual’s viral load is tracked as a function of time since infection (Supplementary Figure S5), along with the viral variant with which an individual is infected (inherited from the infector). The model captures viral transmission as infectious and susceptible individuals come into contact. The probability of transmission is dependent on the viral load of the infectious individual, the variant of the virus being transmitted, and any partial immunity acquired by the susceptible individual (either through previous infection^26^ or vaccination^27, 28^). Further, seasonality affects the probability of transmission (Supplementary Figure S6) - with lower probabilities in warmer periods - reflecting a larger proportion of people coming into contact outdoors once temperatures warm. Any contact between an infectious and susceptible individual is assumed to carry the same probability of transmission, all else being equal. Human contacts are represented through an age-structured network (Supplementary Figure S4)^29^.

A newly infected individual will, following a latent period, be assigned through stochastic distributions an age-dependent disease prognosis of either asymptomatic, mild, severe, critical, or eventual death (Figure 1). These prognosis probabilities are derived from publicly available age-disaggregated morbidity and mortality data^30, 31^. The viral variant an individual is infected with can alter prognosis probabilities, capturing the ability of certain variants to cause increased morbidity and/or mortality (e.g., B.1.1.7^15, 16^). Cases with prognosis of severe, critical, or eventual death may be admitted to hospital following some delay from symptom onset or may alternatively receive care outside of hospital (e.g., in a care home). Critical cases who are in hospital will be admitted to an intensive care unit (ICU), with sufficient capacity assumed in the model. The duration an individual remains in any given disease and/or care state is sampled from a distribution (Supplementary Figure S8).

### Data

Application of OpenCOVID to the national-level epidemic in Switzerland was informed by publicly available demographic and epidemiological data from the Swiss Federal Office of Public Health (FOPH)^32^ and climate data from Meteo Schweiz^33^. NPI measure data from various public sources^25^ were used to compute national- and cantonal-level interpretations of the OHCI. A subset of model parameters (Supplementary Table S1) was calibrated in order to align model output to six types of epidemiological data, including confirmed cases, patients in hospital, deaths, and prevalence of viral variants^10, 13^. A log-likelihood objective function was used to measure the overall quality of the model fit weighted for each data type. The effect of NPIs on contact rates was also subject to model calibration, where the OHCI index was proportionally scaled to represent a reduction in contacts. See Supplementary Information for calibration details and outcomes.

### Diagnosis

Upon infection, an individual is assigned a date at which they may potentially be diagnosed as a consequence of test seeking behaviour. The delay between symptom onset and a potential diagnosis for each individual is sampled from a Gaussian distribution. We assume all non-severe cases isolate for a 10-day period immediately following diagnosis. For individuals presenting with severe disease that seek hospital care prior to diagnosis, a test (and consequent diagnosis) is assumed to be carried out once they are admitted to hospital.

In this application for Switzerland, we derive numbers of diagnoses over time directly from data of confirmed COVID-19 cases. All COVID-19 cases that seek hospital care receive a diagnosis. After taking hospitalised diagnoses into account, other individuals with severe disease outside of the hospital setting and individuals with mild disease are randomly selected and assigned a diagnosis in the model. To represent future test-seeking behaviour, the model-calculated the proportion of cases diagnosed per infected case over the past 14-days and maintained this proportion into the future (Supplementary Figure S9). This assumption is not robust to major changes in testing policies or behaviours, including, but not limited to, mass testing.

### Immunity

Following a period of infection, surviving individuals recover to a state in which viral shedding no longer occurs. These recovered individuals are assumed to be susceptible to reinfection but are assumed to retain some level of partial immunity, which reduces susceptibility to subsequent exposure. OpenCOVID can capture immunity decay, but for this study we optimistically assume no immunity decay following natural infection or vaccination. For naturally acquired immunity due to infection, we assume an 83%^26^ transmission blocking effect when/if re-exposed. If a previously infected but recovered individual is later vaccinated, the level of transmission blocking immunity is taken to be the highest of the two independent effects. No synergistic effect is considered.

### NPIs

In OpenCOVID, non-pharmaceutical interventions (NPIs) can curb the spread of SARS-CoV-2 in an otherwise unprotected population by reducing the number of potentially transmissible pairwise contacts. In Switzerland, NPIs have targeted several aspects of public life, including closure of shops, restaurants and other entertainment venues, restrictions in sizes of spontaneous gatherings, the cancellation of events, and facemask mandates in publicly accessible spaces. The Oxford Health and Containment Index (OCHI) is a measure that is proportional to the amount (or stringency) that such measures are in place at a given moment in time^34^. The level of the OCHI, together with a calibrated multiplicative scaling parameter is used in our model to capture the effect of NPIs in reducing the effective daily number of contacts. The Swiss level of the OCHI is collected on cantonal and federal levels, based on publicly available information^25^. This publicly available information is translated into 16 Swiss-specific variables, and from there into the 13 variables that together make up the Oxford Containment and Health Index.

### Variants

OpenCOVID tracks the transmission of multiple viral variants. Variants are imported into the modelled population 7 days prior to being identified in national genomic surveillance data. We assume the variant with which an individual is infected is inherited from the infector. At the time of writing, B.1.1.7 was the dominant variant in Switzerland, replacing D614G^10, 13^. We modelled three variants in this study: D614G, B.1.1.7, and B.1.351. A 60% increase in B.1.1.7 transmissibility relative to D614G best matched variant prevalence data between January 2021 and February 2021 (Supplementary Figure S3). For B.1.351, a 10% increase in transmissibility relative to D614G was assumed ^12^. For the primary results reported in this study, we assumed no increased probability of morbidity or mortality due to viral variants. However, we assess the impact of this scenario in a sensitivity analysis.

### Vaccines

Fully susceptible, partially susceptible, and actively infected individuals not in hospital can potentially receive a vaccine. Vaccination is modelled using two properties: first, to trigger an immune response that blocks transmission for a proportion of exposure events, and second, to reduce the probability of developing symptoms if infection does occur. Upon vaccination, there is a time delay until the full efficacy of the vaccine is realised; a sigmoidal curve is used to represent this growth in vaccine effect. Vaccine efficacy, delay to full efficacy, transmission blocking effect, and number of doses required is vaccine specific (Supplementary Table S5). In this study, we consider only mRNA vaccines and assume all vaccinated individuals receive two doses spaced by 3-weeks with vaccination reaching maximum efficacy 28-days after the first dose^27, 28, 35^. We assume the vaccine is 80% transmission blocking and has a further 75% probability of preventing symptoms leading to the observed 95% vaccine efficacy reported in clinical trials^27, 28^. In this study, we do not model decay of vaccine efficacy over time or reduction in vaccine efficacy due to variants of concern, but the model is able to capture changes to these assumptions.

### Model simulations

This analysis and COVID-19 model outcomes are conducted at the national level, but this model can also be applied at the subnational level. All outcomes are reported as mean estimates alongside prediction intervals representing parameter and stochastic uncertainty. Model simulations were initiated on 18 February 2020, 7-days before the first cases were confirmed for three consecutive days (25 to 28 February 2020) in Switzerland. All model processes were computed at 1-day time intervals. A number of initial cases were imported into the population, which were then able to cause onwards infection. A number of infections are also imported into the population at each time step.

Furthermore, new virus variants are initiated by importing a number of new cases of each variant at a given time in alignment with the point the particular variant was first identified in Switzerland. All three importation rates are found through model calibration (see the following section). The number of individuals simulated in the model is capped at a predefined number (one million individuals for all simulations), with a population scaling factor applied to all relevant model outputs to represent a one-to-one scale for the Swiss population of 8.5 million.

Numerous model outputs are captured and reported temporally, including number of infections, diagnosed infections, morbidity, and mortality estimates (Supplementary Figure S1). Where appropriate, metrics are disaggregated by age and variant of concern (Supplementary Figures S2 and S3).

### Scenarios

The OpenCOVID model was used to predict future national-level epidemic trajectories from early March to early September 2021. We did not simulate beyond this date due to substantial uncertainty around duration of natural and vaccine-induced immunity, the impact of additional new variants, and changes in NPIs, adherence, and testing.

Two vaccination roll-out speeds (slower, 50,000 doses per day, and faster, 100,000 doses per day) and a range of NPI relaxations were modelled. Vaccine rollout up to 31 March 2021 was modelled as per publicly available data^36^. Vaccine eligible individuals were assigned to one of five priority groups according to age, comorbidity, and profession, and vaccines were not given to hesitant individuals. Priority groups were modelled with subsequent vaccination with two doses of an mRNA vaccine following the Swiss FOPH strategy^37^ (see Table 2 for further details). As per current Swiss vaccination guidelines, individuals under 18 years of age are not considered eligible for vaccination^37^. We assumed that 75% of each priority group was willing to be vaccinated. NPI relaxation scenarios were modelled as described in Table 1. An NPI ‘relaxation step’ translates to approximately 5 points on the OHCI, reflecting the set of NPI relaxations (further described in Supplementary Information) proposed by the Swiss Federal Council to occur initially on 1 April 2021 (subsequently re-scheduled to 22 March 2021^38^). For all scenarios we assumed adherence to measures was consistent over time. Future scenarios assume similar levels of testing to that from early February 2021 to early March 2021, the potential impact of mass testing or widespread testing outreach was not modelled.

### Sensitivity analysis

To assess the sensitivity of our findings, we simulated scenarios that independently varied several key model parameters related to vaccine characteristics and currently circulating variants of concern (see Table 2 for details).

## Data Availability

The model code is open source and available here: https://github.com/SwissTPH/OpenCOVID
Epidemiological data and demographic data informing the models was obtained from publically available sources

## Acknowledgments

We would like to thank the unit members of the Disease Modelling Unit, Swiss Tropical and Public Health Institute, and also Dylan Muir for insightful feedback on the modelling and manuscript. We would also like to thank Martin Ackermann, Tanja Stadler, Olivia Keiser, Thomas Van Boeckel, Jacques Fellay, Richard Neher, Emma Hodcraft, Jan-Egbert Sturm, Marius Brülhart, Suzanne Suggs, and Nicola Low for their feedback and discussion on this work. Calculations were performed at sciCORE (http://scicore.unibas.ch/) scientific computing core facility at University of Basel.

Funding was provided by the Botnar Research Centre for Child Health (to MAP) and the Swiss National Science Foundation NRP 78 Covid-19 2020 (4078P0_198428). MAP is funded by a Swiss National Science Foundation Professorship (PP00P3_17070).

This work has been possible thanks to many members of Swiss National COVID-19 Science Taskforce and Swiss Federal Office of Public Health, who contributed discussion and supported model scenarios and interpretations.

## Contributions

MAP, NC, and AJS conceived the study. AJS and RPD developed the model with input from ELR, SS, and SLK. Analyses were performed by AJS, EALR, and RPD. AJS and MAP validated the model and analyses. Figure preparation was performed by AJS and EALR. All authors contributed to interpretation of the results, writing the draft and final version of the manuscript, and gave final approval for publication.

## Competing Interests statement

The authors declare no competing interests.

## Code availability

The code used for this analysis is available from https://github.com/SwissTPH/OpenCOVID

## Supplementary Information

### Model calibration

OpenCOVID was calibrated to the national-level epidemic in Switzerland using publicly available epidemiological data from the Swiss Federal Office of Public Health (FOPH) [1]. Model output was matched to six types of observed temporal metrics: 1) daily confirmed COVID-19 cases, 2) daily COVID-19-related deaths, 3) daily hospital admissions, 4) number of COVID-19 patients in hospital, 5) number of COVID-19 patients in ICU, and 6) relative prevalence of virus variants [2, 3]. Figure S1 shows the alignment of the model output to the epidemiological data, along with several additional model metrics. Age-disaggregated metrics are illustrated in Figure S2. The alignment of the model to data regarding prevalence of viral variants is shown in Figure S3. See Table S1 for a full list of calibrated and fixed model parameters.

### Parameter table

### Variants of concern

OpenCOVID tracks transmission chains of viral variants. The model can consider any number of variants, providing there is sufficient data to inform the relative prevalence of each variant in the population. Three viral variants were modelled in this application of Switzerland: D614G (considered the dominant variant at the start of the epidemic), B.1.1.7, and S501Y V2. Figure S3 shows the alignment of the calibrated model to variant prevalence over time. We modelled the B.1.1.7 and S501Y V2 variants by assigning them a percentage increase in the probability of transmission per contact then further calculated the likely transmission advantage in a heterogeneous population with pre-existing immunity during an ongoing pandemic with existing non-pharmaceutical interventions (captured in our individual-based model). We estimated the effective reproductive number, Re, of B.1.1.7 for the months of January and February, for the specified increase in the transmission probability, assuming a serial interval of 6-days and calculated the transmission advantage as the ratio of the effective reproductive number to that of variant D614G. We further conducted a sensitivity analysis varying the serial interval (3-9 days).

The transmission advantage is therefore the proportional increase in the expected number of cases from one infected individual in the epidemic setting in Switzerland in early 2021 (including the effects of pre-existing natural immunity and the impact of control measures). It is thus important to note that a 60% increase in the probability of transmission per contact does not correspond to a 60% increase in the effective reproductive number, Re, but rather closer to a 30% increase. Allowing for variation in the serial interval, this can vary between 10% and 50%. The scenario with a 70% increase in transmissibility corresponded to a transmission advantage of 1.4 (1.2-1.6); and that with a 50% increase in transmissibility corresponded to a transmission advantage of 1.2 (1.1-1.4).

### Likelihood and calibration details

The calibration process for this application to Switzerland was as follows: 4,000 parameter sets were initially sampled across parameter hyperspace. That is, the region defined by the bounds of each calibrated parameter in Table S1 (calibrated parameters highlighted in blue). A Latin hypercube algorithm was used for this initial sampling; thus, no focus was placed on regions in which parameter priors are located. Each parameter set was then simulated 10 times for different stochastic realisations using OpenCOVID, thus capturing a basic understanding of stochastic uncertainty. All model simulations were performed simultaneously on a high-performance computing cluster [33]. The value of a log-likelihood objective function was calculated for each parameter set, quantifying the likelihood of the model parameters given the epidemiological data illustrated in Figure S1 and Figure S3. The log-likelihood function is given by:

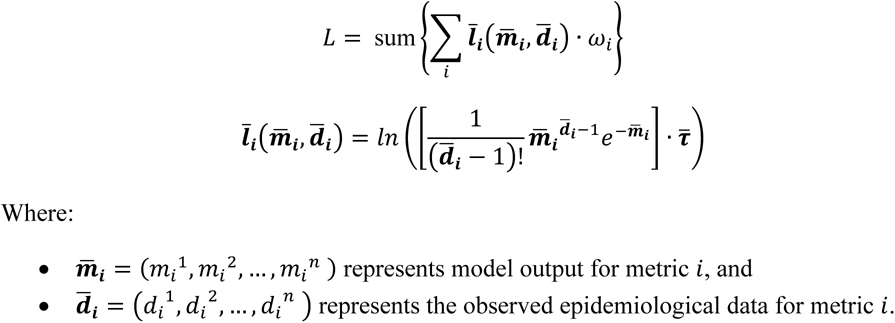

The values *m_i_*^1^ and *d_i_*^1^ represent the first date for which we have non-zero data for metric *i*, starting from 24^th^ February 2020, whilst*m_i_*^n^ and *d_i_*^n^ represent the final date for which we use data in the model calibration process. In this application of Switzerland, *n* represents 5^th^ March 2021. The vector 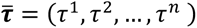 is the time weight vector, where 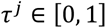 for all *j*. In this application 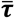 is defined to linearly increase from 0.5 to 1 between 24^th^ February 2020 and 5^th^ March 2021. The constant *ω_i_* is the weighting applied to metric *i*. These weightings used for this application are shown in Table S3.

A Gaussian Process model was then trained using all parameter sets and associated likelihood values to predict the log-likelihood over all data given a model parameter set. For this analysis, we used a heteroscedastic Gaussian Process algorithm as the model emulator [34, 35]. Ten rounds of adaptive sampling were applied to efficiently resample regions of the parameter hyperspace that were good candidates for the global maximum. An expected improvement acquisition function was used to identify these candidate regions and sample 100 new parameters sets per round, with a filtering function applied to ensure resampled parameters sets are not within a predefined distance of each other (with distance measured in Manhattan space). Following the ten rounds of adaptive resampling, the simulated parameter set with the highest log-likelihood value (considering the mean over all stochastic simulations) was identified as the best estimate parameter set, as reported in Table S1. See Figure S1 and Figure S3 for alignment of model outcomes to the observed data.

### Infectiousness per contact

In OpenCOVID, we define a pairwise transmissible contact to be a human-to-human contact that has a transmission probability of β when the infectious individual is *fully infectious,* and the susceptible individual is *fully susceptible*. An individual is *fully infectious* when their viral load is at a maximum (see *Viral load profile* section). An individual is considered to be *fully susceptible* when they have zero immunity (see *Immunity* section). We note here that in this application, both previously infected and vaccinated individuals will possess a non-zero level of immunity. Two additional factors can alter the probability of transmission between and infectious individual and a susceptible individual. First, a seasonality effect reduces the probability of transmission in warmer periods, reflecting a larger proportion of contacts being outdoors with warmer temperatures (see Seasonality section). Second, novel viral variants can enter the population, being more (or less) infectious than the current dominant variant, and therefore increase (or decrease) the probability of transmission. For this study, we assume the SARS-CoV-2 epidemic in Switzerland began with variant D614G being the dominant variant (see Figure S3). For this study, we defined β to be 5% (see Table S1). That is, we define a contact to have a 5% probability of transmission when the infectious person has a peak in viral load, the susceptible person has no partial immunity, the contact is during the coldest day of the year, and the variant being transmitted in D614G. With the value of β fixed, the population average number of contacts can then be calibrated such that observed epidemiological data is matched (see *Contact network*, *Model calibration*, and *Parameter table* sections).

In equation form, the probability of transmission between an infection individual, *I*, and a susceptible individual, *S*, is given by:

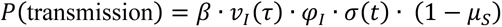

Where:

- *ν*_I_(τ)ε[0,1] denotes the viral load of the infectious individual, *τ* days following infection (see *Viral load profile* section).
- φ*_I_* denotes the infectivity factor of the viral variant with which the infectious individual is infected (see
- Table S2 in *Variants of concern* section).
- σ(*t*) denotes the seasonality scaler at date *t* (see *Seasonality* section)
- *μ*_S_ denotes the immunity of the susceptible individual (see *Immunity* and *Vaccine properties* sections)

### Contact network

The contact network in OpenCOVID is based on the POLYMOD contact survey [36] which reports age-structured contact frequencies. The POLYMOD survey is implemented in OpenCOVID via the R package *socialmixr* [37] which provides symmetric matrices in which the rows and columns are the age class of the ego (the person reporting the contact) and the alter (the person receiving the contact), and the cell content is the average number of contacts between those age classes. This data can be accessed by country. As POLYMOD does not provide Swiss survey data, we use artificial contact frequencies based on survey data from France, Germany, and Italy. We then use Swiss age-structured demographic data to sample this contact frequency space and create an age-structured random network by sampling with replacement, weighted by the average number of contacts per cell. We sample such that the resulting network has a mean number of contacts (as defined by the ‘contacts’ parameters, see Table S1). In such a network, not all age classes have the same number of contacts. Younger age classes have more contacts and especially have more contacts with other young age classes while older age classes have fewer contacts. See Figure S4 for an illustration. This captures the qualitative aspect of an age structured network based on European survey data, and then transforms to Swiss specific demography. This network does not distinguish between work, school or home networks but is rather integrated across all these separate networks. Individual ages are tracked for 0-90 years in one-year age bins, with an additional group for 90+. Gender is not considered in the model.

With *β* (the infectiousness per contact, see Infectiousness per contact section) set at a fixed value (see Table S1), the population average number of contacts can then be calibrated such that observed epidemiological data is matched. The primary signal for the contacts parameter is the exponential increase in all observed metrics during the first wave prior to the observed impact of NPIs.

### Viral load profile

During the latent period that follows infection, we assume viral load is zero (and therefore that the infected person is not yet infectious). We then use a gamma probability density function with shape parameter *α* = 3 and rate parameter β = 0.5 to represent individual-level viral load over the course of the infectious period. We assume infectiousness is proportional to this viral load, and therefore standardise viral load values to between zero and one to convert viral load into an infectiousness scaler that scales the probability that the individual can infect other contacts. The parameters of the gamma function were selected to best represent the current understanding of viral load profiles from time since infection [38, 39]. Figure S5 illustrates this infectiousness scaler profile from time since infection. In equation form, the infectiousness scaler for an individual *k* infected τ days after infection is given by

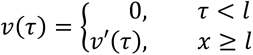

where *l* is the sampled latent period for individual *k* (see ‘Infection, disease, and hospitalisation durations’ for duration distributions) and

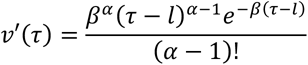

### Seasonality

We take daily temperature data from the federal office of meteorology and climatology MeteoSwiss [40] which is made available via the opendata.swiss service [41]. We use the daily maximum temperature values that are available from the NBCN measurement station network [42]. These stations provide different daily weather data across Switzerland. Nevertheless, not all cantons have stations, and some cantons have several stations. For national level analyses we use averaged data across all available weather stations. For cantons with multiple weather stations, we take the mean value across all stations. We then use a population-weighted mean to derive national level values. In Figure S6, the grey lines represent temperature data from 1^st^ January and 5^th^ March 2021 (date of calibration). Over this time period, the yellow line represents the population-weighted national average. From 6^th^ March onwards, the line represents the future projection of temperature. This future projection uses monthly data from years 1981-2010, and is generated by a spline fitting algorithm form the R package *RMAWGEN* [43]. The seasonality effect used in the model (see *Infectiousness per contact* section) is then derived from the normalized inverse of the temperature curve.

### Non-pharmaceutical interventions

In OpenCOVID non-pharmaceutical interventions (NPIs) can curb the spread of SARS-CoV- 2 by reducing the number of potentially transmissible pairwise contacts. In Switzerland, such measures have included the closing of non-essential shops, restrictions on mass gatherings, and facemask mandates in publicly accessible spaces. The Oxford Containment and Health Index (OCHI) is a measure that is proportional to the amount (or stringency) of such measures that are in place at a moment in time [44, 45]. The OCHI ranges from 0 to 100, with 0 being no measures in place and 100 being the most restrictive full lockdown possible. The level of the OCHI, together with a calibrated multiplicative scaling parameter is used in OpenCOVID to capture the effect of NPI in reducing the effective daily number of contacts. In effect, the *edge list* associated with the contact network – that is, the list of all daily pairwise contacts in the network – is reduced by a proportion given by the product of the OCHI on a given day and the calibrated NPI scaling constant (see Table S1). In this manner, we do not explicitly simulate the effect of individual measures, but instead model the total effect of all NPIs in place.

The Swiss level of the OCHI is collected at the cantonal- and federal-level based on publicly-available information from various sources, and is available from the SwissTPH GitHub [46]. This publicly available information is translated into 16 Swiss specific variables, and from there into the 12 variables that together make up the Oxford Containment and Health Index. The SwissTPH GitHub on the Swiss measures provides a codebook for the Swiss specific variables. For an overview of the variables that make up the Oxford Containment and Health Index, their coding schemes and how to calculated the OCHI from the constituent variables, the reader is referred to the codebook, the coding interpretation guide, and the instruction on how to calculate indexes provided by the Blavatnik School of Government at the University of Oxford [44, 45].

Figure S7 shows the value of the Oxford Containment and Health index, that is representative of the strength of measures as applied in Switzerland, for past data (left of the vertical black dashed line) as well as an example scenario of future NPI relaxation (right of the vertical black dashed line) which corresponds to the red scenario from figure 2 in the main manuscript. This scenario considers three different five point NPI relaxation steps, from 58.5 to 53.5 on 22 March 2021, to 48.5 on 12 April, and to 43.5 on 5 May. The five percentage point relaxation steps were chosen to approximately represent a potential NPI relaxation package published by the Federal Council on 17 February 2021 [47]. This potential package included increasing the limit on indoor private events from 5 to 10, opening professional sporting and cultural events at one-third capacity, and reopening restaurants for outside service [47]. It is important to note that not all openings have a quantitative effect on the OCHI. We stress here that the specific openings are not modelled explicitly, but rather the abstraction of an equivalent amount of NPI relaxation that is reflected in the OCHI. Moreover, the same amount of opening in terms of OCHI can also be reached with different combinations of openings, so that the OCHI stays agnostic to a specific type of opening, and only reflects a certain amount of opening. We provide all gathered detailed information on the measures that were in place at all dates, both at the cantonal and national level on the Swiss TPH GitHub [46].

### Prognosis probabilities

Once infected with SARS-CoV-2 and following a latent period, an infected individual develops either asymptomatic, mild, or severe disease. Individuals that develop severe disease may, after some time, either seek hospital care or remain outside of the hospital setting (e.g., within care homes). We model three distinct prognosis tracks for those that will seek hospital care: 1) the patient will eventually recover without intensive care, 2) the patient will require intensive care but will eventually recover, and 3) the patient will require intensive care and will ultimately die from COVID-19-related complications. See manuscript Figure 1 for an illustration of modelled natural history and prognosis pathways. We quantify age-group stratified probabilities for each prognosis using publicly available age-disaggregated morbidity and mortality data [47, 48], see Table S4. Once infected, a prognosis is derived for all individuals by stochastically sampling from a uniform distribution.

The prognosis probabilities given in Table S4 assume an equal probability of infection across all age groups. Whilst the probability of infection in any given contact is not assumed to be age-dependent (see *Infectiousness per contact* section), the number of contacts for any given person *is* age-dependant (see Figure S4 in *Contact network* section). Therefore, each age-dependent prognosis probability needs to be scaled by an age-correction factor to convert to per-infection probabilities. Three additional factors can affect these age-related prognosis probabilities:

1. Improved care procedures (see improved_care_factor, Table S1)
2. Increased mortality of viral variant infected with (see Table S2)
3. Symptom reducing effect of vaccination

The *improved care* factor represents the reduction in hospitalized COVID-19 cases requiring intensive care due to improved triage, use of treatments such as dexamethasone, and other factors. This improved care factor is calibrated, and assumed to take an effect on 1^st^ June 2020 following the ‘first wave’ experienced in Switzerland. For individuals infected following vaccination, an age-dependent prognosis is initially derived as described above. A further probability of being asymptomatic instead of symptomatic is then calculated by multiplying the *symptom reducing effect* of the vaccine with the normalized level of vaccine efficacy at that point in time (see *Vaccine properties* section).

### Infection, disease, and hospitalisation durations

Upon infection, the duration for which an individual will remain in each disease or care state is sampled from a distribution, as illustrated in Figure S8, and described in Table S1 (including sources for the best estimated values for each duration).

### Testing, diagnosis, and isolation

Upon infection, an individual is assigned a date at which they may potentially seek a test and be diagnosed as a confirmed COVID-19 case. The delay between symptom onset and a potential diagnosis for each individual is sampled from a truncated Gaussian distribution (see Figure S8). We derive the number of diagnoses over time directly from observed data of confirmed COVID-19 cases (see Figure S1) and apply the relevant number of diagnoses per day across the modelled population. By definition, all COVID-19 cases that seek hospital care receive a diagnosis. After taking hospitalised diagnoses into account, other individuals with severe disease outside of the hospital setting and individuals with mild disease are randomly selected as those who seek testing and are assigned a diagnosis in the model. To represent future test-seeking behaviour, the model-calculated proportion of cases diagnosed per infected case over the past 14-days is fixed into the future (Figure S9). We note here that this assumption is not robust to major changes in testing policies or behaviours, including, but not limited to, mass testing. We assume no change in behaviour for individuals who test negative, and further assume that all non-severe cases isolate for a 10-day period immediately following diagnosis.

### Immunity

For individuals that recover from SARS-CoV-2, we assume a partial acquired immunity of 83% to future infection upon recovery regardless of disease severity, risk group, or age [49-51]. For the relatively short-term projections presented in this application to Switzerland, we assumed no waning of acquired immunity. We note here that this optimistic assumption may not be appropriate for longer-term projections. In future work, the assumptions regarding level of immunity by disease state and waning acquired immunity will likely be reassessed as new evidence becomes available.

### Vaccine properties

Vaccine efficacy following two doses was assumed to depend on vaccine type. For the mRNA vaccines (Pfizer and Moderna), an efficacy of 95% was assumed for all priority groups P1– P5. We implemented the vaccination of an individual as the smooth increase in immunity over time using a sigmoidal function that has a lower asymptote of 0, an upper asymptote of overall ‘vaccine efficacy’ (95% for Pfizer and Moderna, 62% for AstraZeneca) and an inflection point 14-days after vaccination. The growth rate of this curve is such that vaccine efficacy is close to zero on the day of vaccination, and is closer to full ‘vaccine efficacy’ after 28-days (see Figure S11).

Most vaccine trials claim to reduce COVID-19 symptom development as well as disease severity, however it remains unclear to what extent they prevent transmission. As a baseline, we assume mRNA vaccines are 80% effective in preventing infection (and future transmission) when at full efficacy. We then calculate the additional effectiveness of the vaccine to reduce symptoms such that the total reduction in symptoms among those vaccinated when the vaccine is at full efficacy (that is, 95% from 28 days after receiving the first dose).

By definition we have that

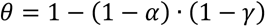

Where θ is the overall efficacy of the vaccine in symptomatic COVID-19, α is the transmission blocking effect, and γ is the additional symptom reducing effect. Solving for *γ*, we have

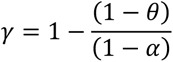

In this application, we assess the sensitivity of model outputs to different assumptions of the vaccine fully protecting from infection (sterile immunity and preventing onward transmission). Namely, values of 60% and 95% for sterile immunity. The corresponding additional symptom reducing effect in each case is reported in Table S5.

### Vaccine rollout

Vaccination strategies were modelled according to FOPH priority groups and updated (10 February 2021) after input from FOPH and other experts to reflect current rollout realisation (Table S6). We assumed a baseline coverage of 75% across all priority groups for most scenarios. For scenarios 4A) and 4B) we modelled 60% and 90% coverage for priority groups P2 – P5 as a sensitivity analysis. We assumed either 50,000 or 100,000 vaccine doses used per day from 1 April 2021, which is within the bounds of the maximum vaccine availability as expected by the FOPH. Until 5 March we use data for number of doses used as provided by FOPH and scale linearly from there up to the target daily doses by 1 April. With 100,000 doses per day the target coverage of 75% across all groups will be reached before July 2021, with 50,000 doses per day it will not be reached before the end of the simulation in September 2021. OpenCOVID vaccinates people strictly according to priority groups, with the highest priority group receiving all doses until the target coverage is reached. Vaccines from CureVac, Novavax, and AstraZeneca were not incorporated in our projections.

### Simulation details

Model simulations were initiated on 18th February 2020, 7-days before the first cases were confirmed for three consecutive days in Switzerland (25th to 28th February 2020). All model processes were computed at time intervals representing one day. One million individuals were modelled for simulations reported here, with a population scaling factor subsequently applied to all relevant model outputs to represent a one-to-one scale for the Swiss population. Where appropriate, metrics were disaggregated by age, variant of infection, and vaccine priority group. All model simulations were performed at sciCORE (http://scicore.unibas.ch/) maintained by the Scientific Computing Center at University of Basel.

### Model development and maintenance

OpenCOVID is written primarily in the R programming language [52] and is stable with R version 3.6.0. The code for OpenCOVID is open source, and available from the Swiss TPH GitHub [53]. Due to the level of computational power required to calibrate the model and simulate scenarios, the model pipeline makes use of a SLURM based cluster sciCORE (http://scicore.unibas.ch/) maintained by the Scientific Computing Center at the University of Basel. Interactions to the cluster are written in bash script. The authors of the manuscript maintain the model source code.

---

**Figure S1:**
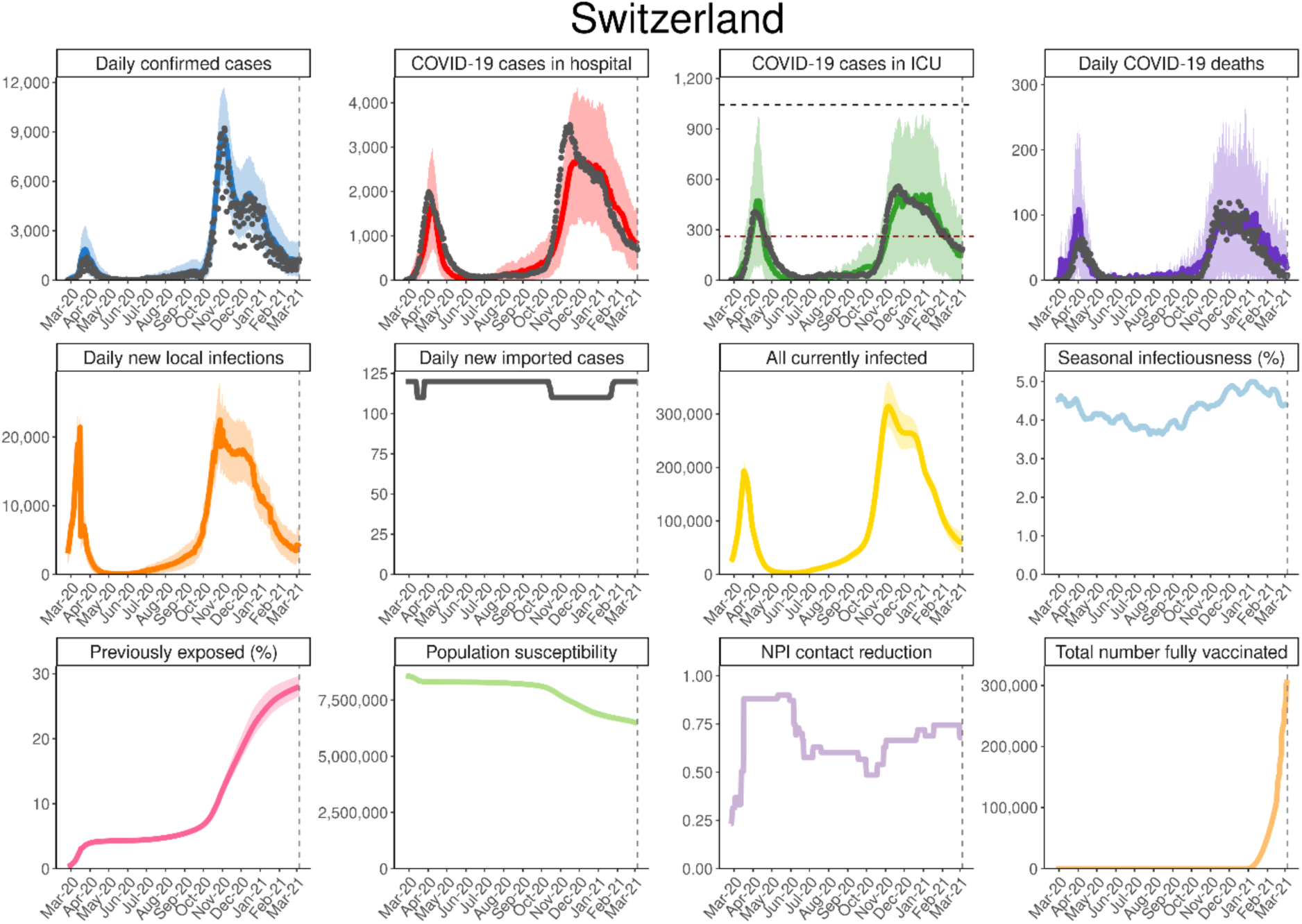
OpenCOVID calibrated to national-level data of confirmed cases, hospitalisations, ICU occupancy, and deaths in Switzerland up to 5 March 2021. Black dots represent the data to which the model has been calibrated. Coloured lines represent model output. Daily hospitalisation rates are not shown due to discrepancies in the data (as described in more detail in the *Likelihood and calibration details* section).

**Figure S2:**
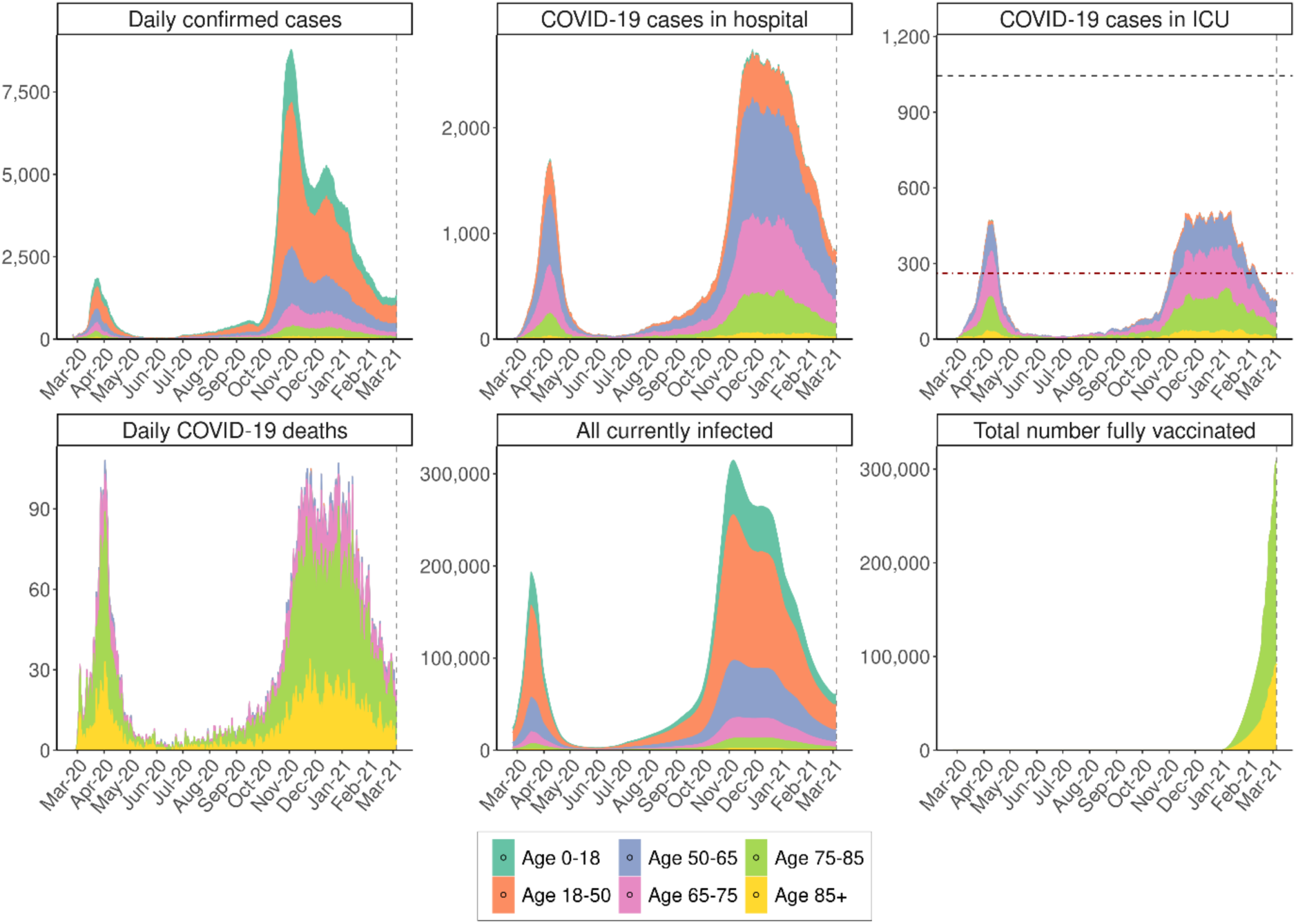
Daily confirmed cases, COVID-19 cases in hospital, COVID-19 cases in ICU, daily COVID-19 deaths, all currently infected and total number fully vaccinated split by age group. Outputs are shown for the calibrated period between 18 February 2020 and 5 March 2021.

**Figure S3:**
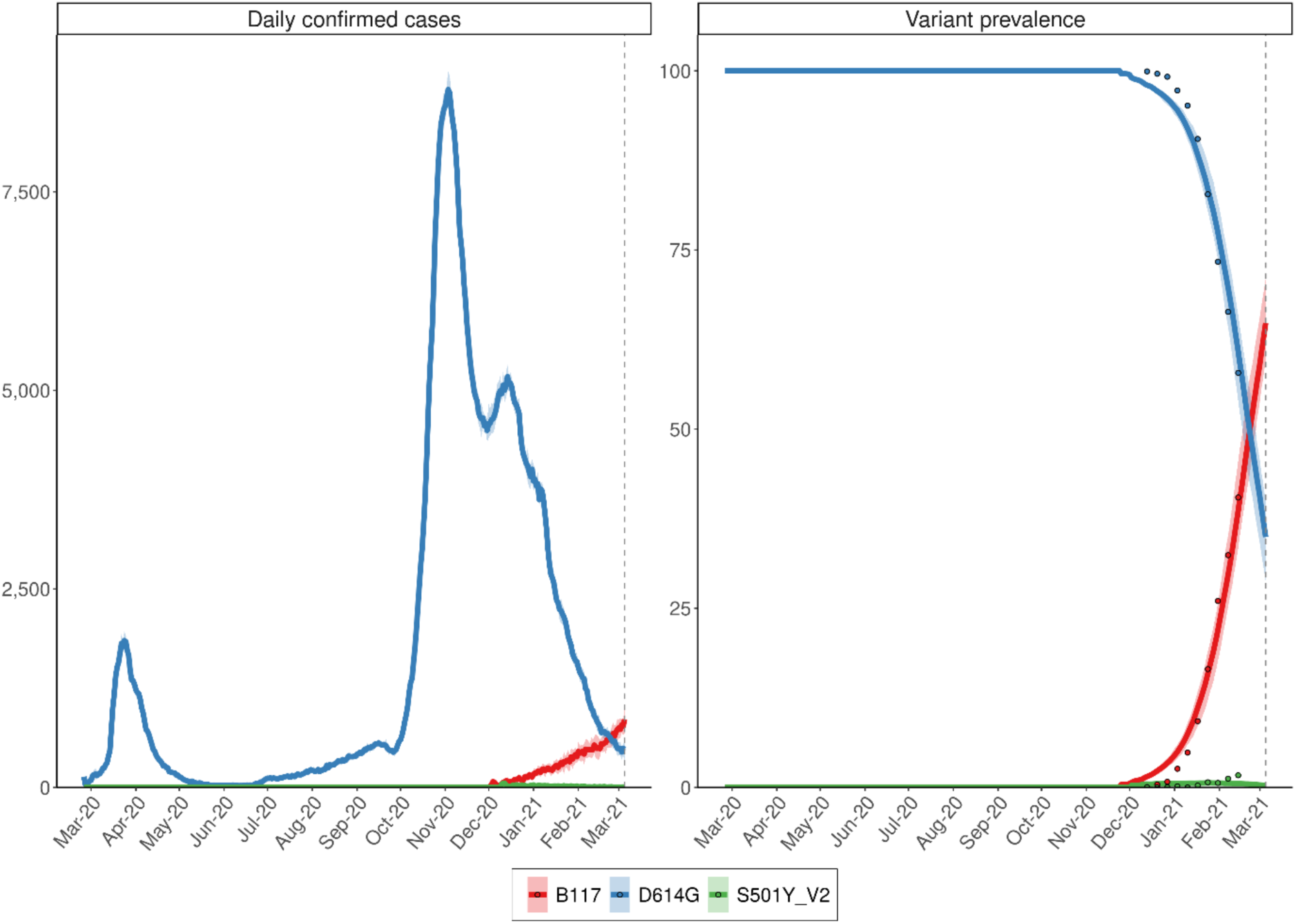
Calibrated variant dynamics in Switzerland until 5 March 2021. Blue is the D614G variant which was dominant in Switzerland from summer 2020 on and is used here synonymously with the earlier strains circulating in Switzerland due to their epidemiological similarity. Red is the B.1.1.7 variant with a 60% increased infectivity. Green is S501Y V2 with a 10% increased infectivity. The solid lines are the model fit, the coloured points is the data.

**Figure S4:**
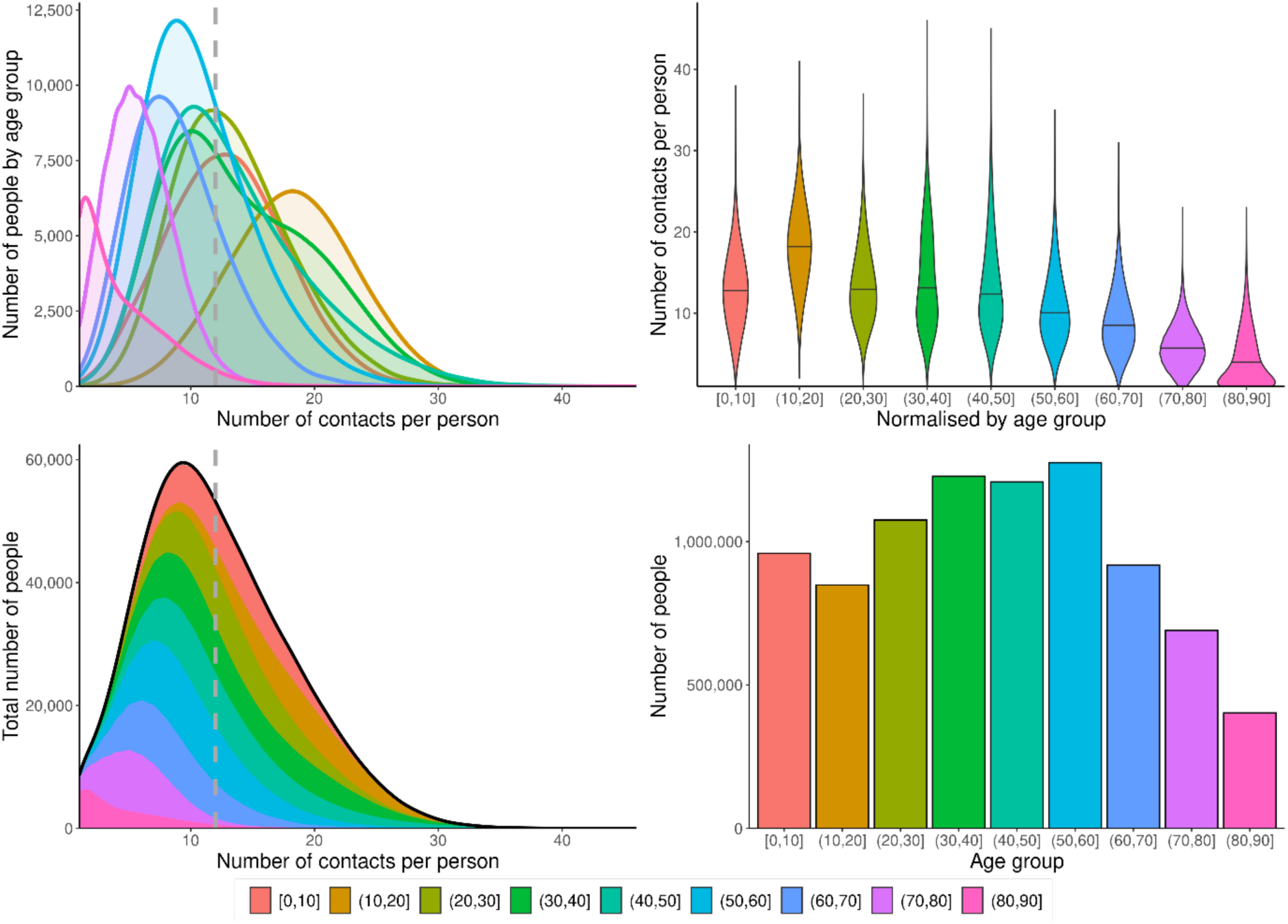
Age-related contact properties in OpenCOVID for this application to Switzerland. Top: Number of people in age group vs. number of contacts per person. Shows both the groups’ sizes and the distribution of contacts. Note that younger people have more contacts. Top right: Normalized number of contacts per person vs. age group. Again, shows the distribution of the average number of contacts per person in an age group. The age group of 10 to 20 year olds has the highest number of contacts. Bottom left: Total number of people vs. number of contacts per person. Bottom right: Number of people vs. age group. Shows the distribution of age group sizes.

**Figure S5:**
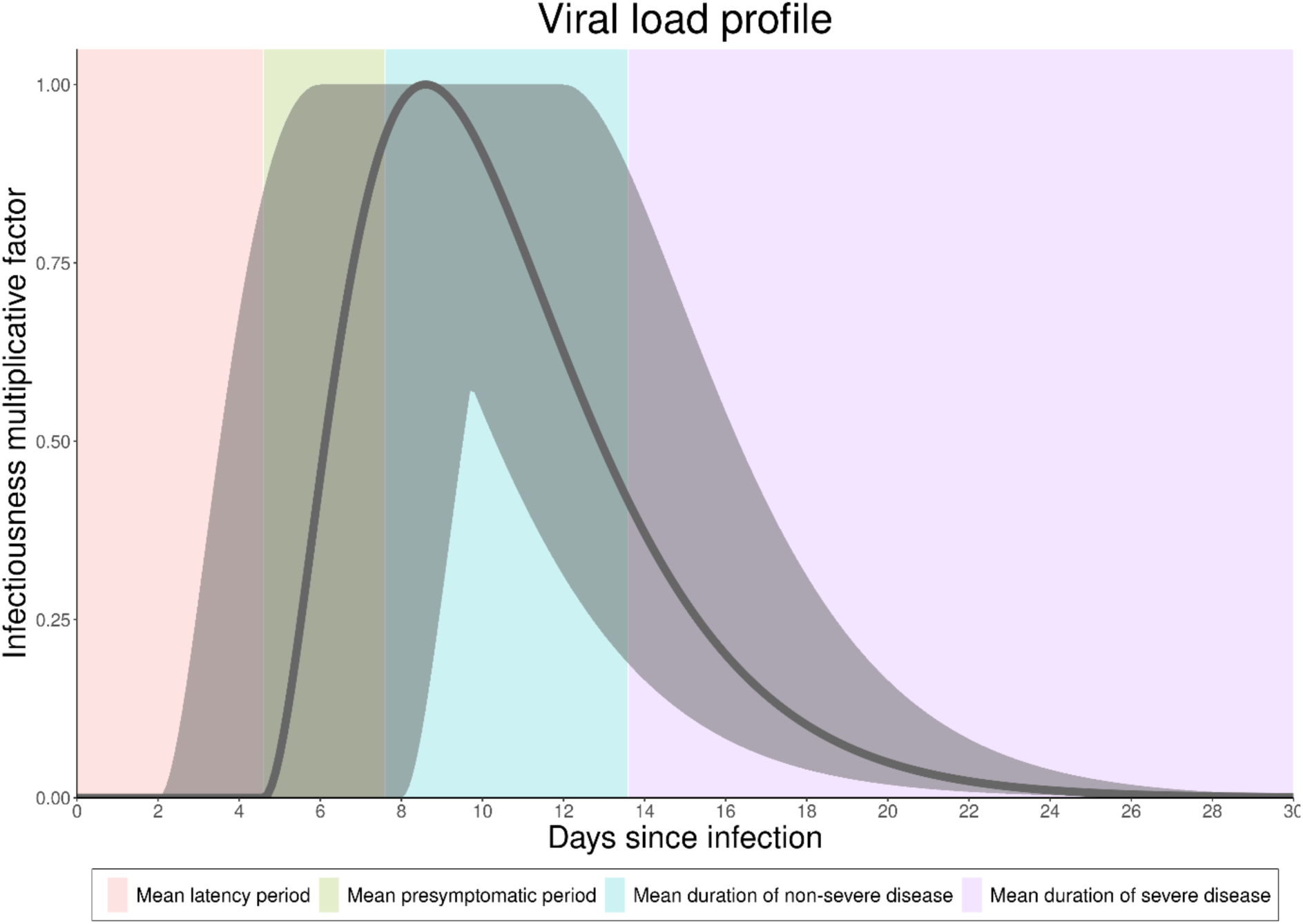
Viral load profile from time since infection. This curve is standardised to between zero and one to result in an infeciousness multiplier used to calculate the probability of transmission. Peak infectivity is reached between days 6 and 14 following infection.

**Figure S6:**
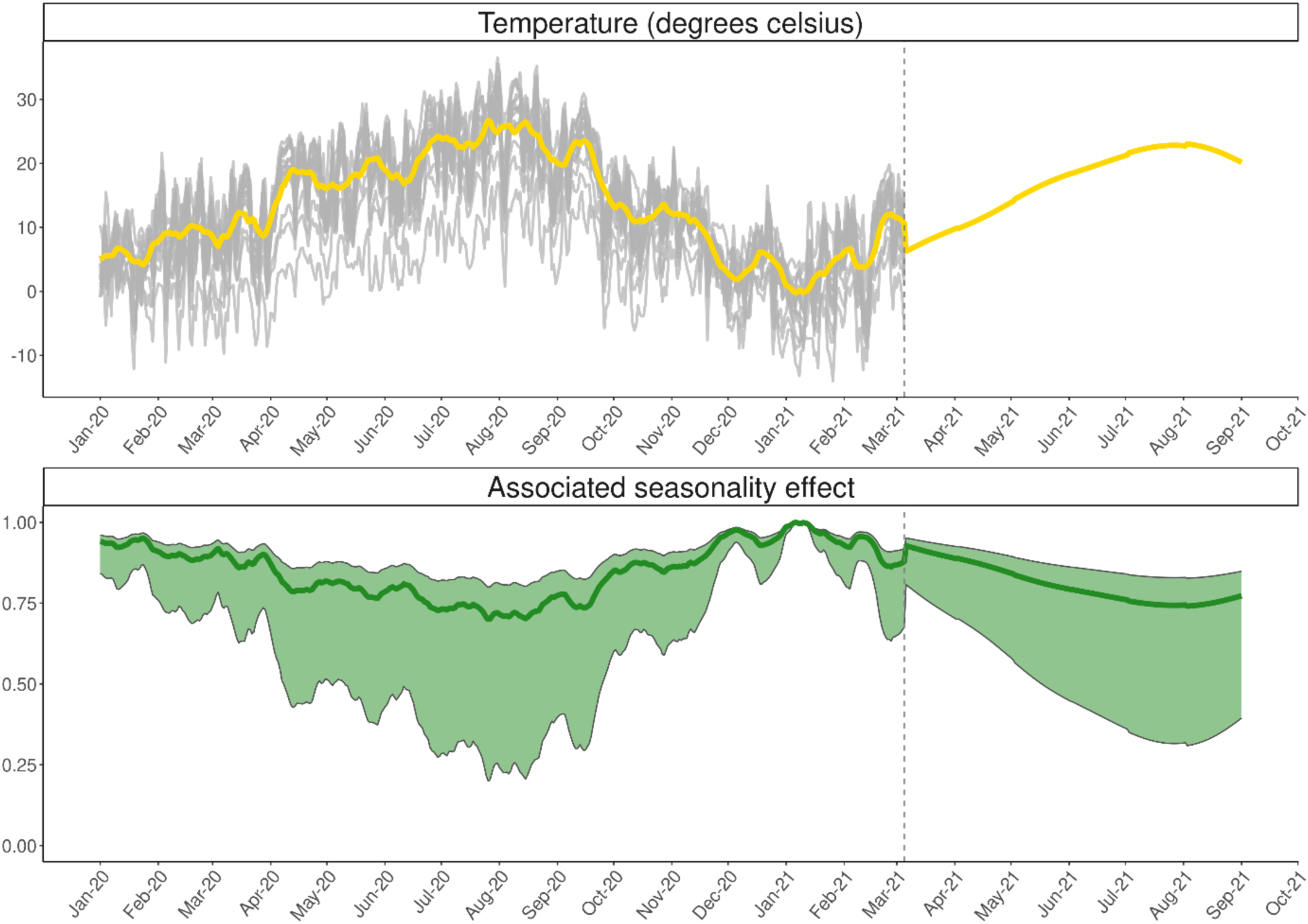
Temperature and associated seasonality effect in OpenCOVID. Grey lines represent past cantonal temperatures, and the yellow line represents the population-weighted national average. The green line represents the associated best estimate seasonality effect. That is, with a seasonality scaler of 0.27 (as reported in Table S1). The green shading represents the range of possible seasonality effects, considering the bounds of the seasonality scaler (see Table S1).

**Figure S7:**
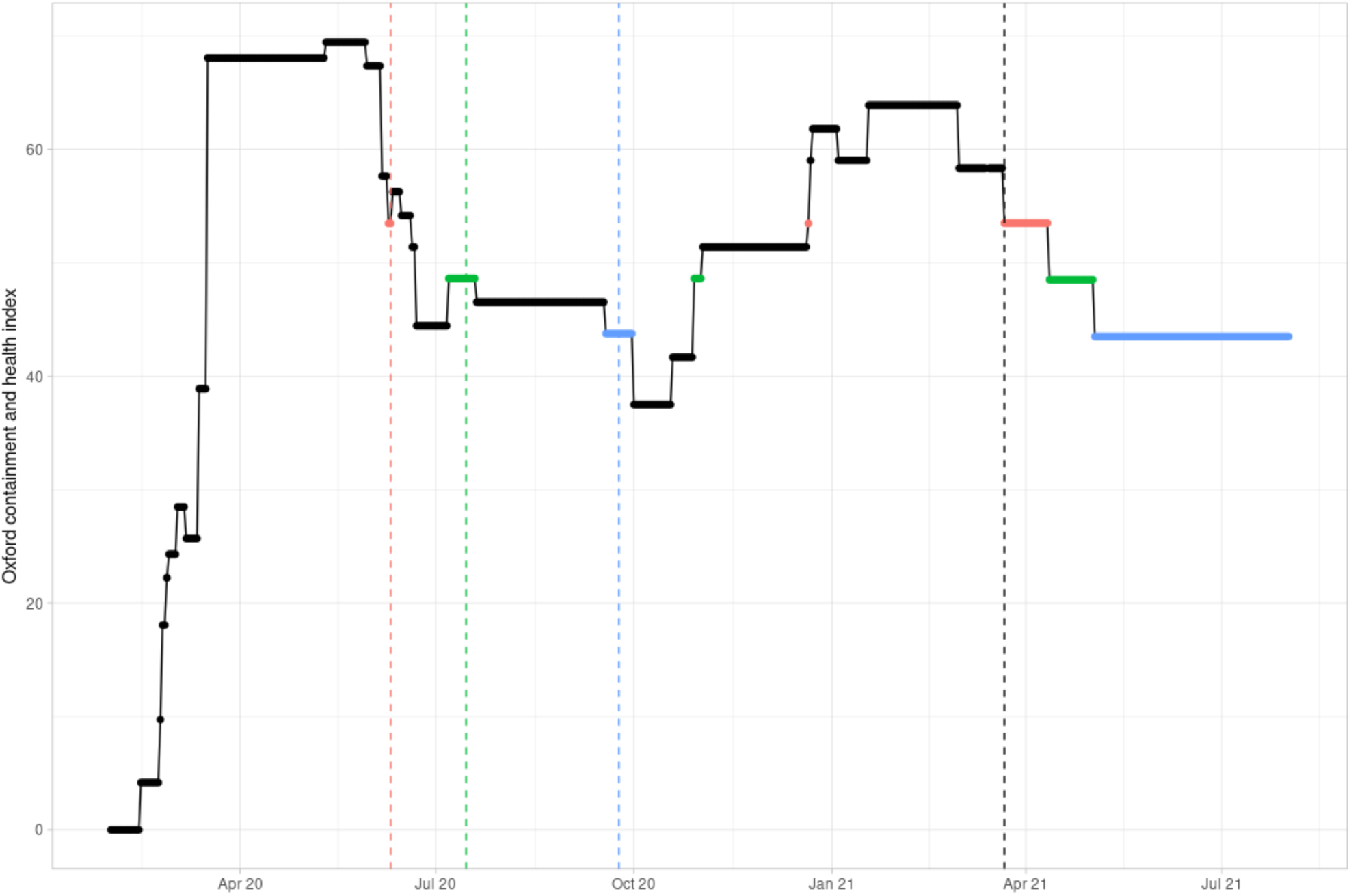
Non-pharmaceutical interventions (NPIs) as Oxford Containment and Health Index over time in Switzerland as implemented at the national level. The depicted NPI relaxation scenario corresponds to the red NPI relaxation scenario from figure 2 in the main manuscript. The red scenario represents five NPI relaxation steps, from 58.5 to 53.5 on 22 March, to 48.5 on 12 April and to 43.5 on 5 May 2021. The vertical dashed black line represents 22 March 2021, the date of the first potential NPI release that was simulated. The colours of the future scenario and the coloured vertical lines highlight the dates when previous NPIs were at a similar level to the future opening steps. Higher values depict stricter measures.

**Figure S8:**
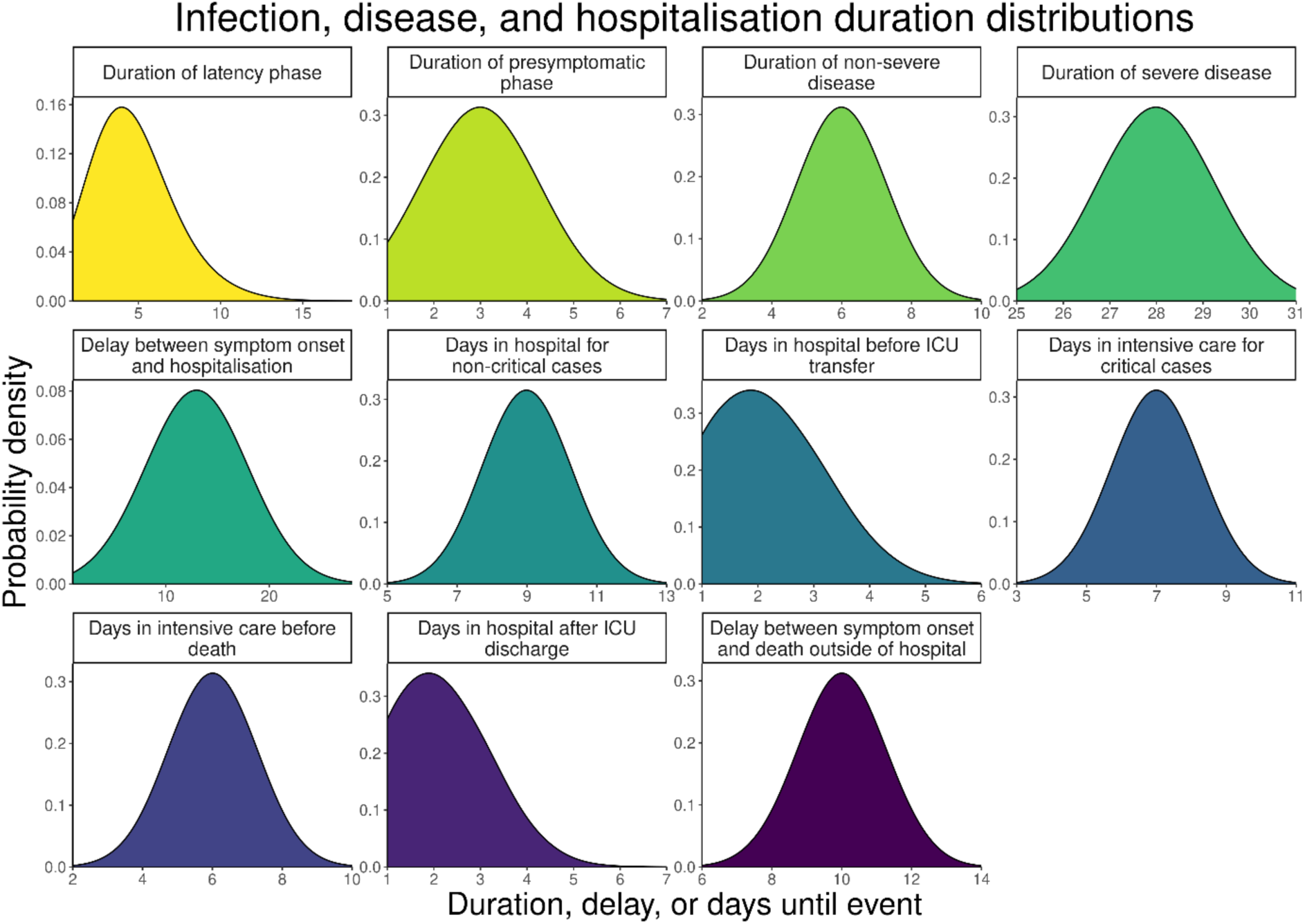
Distributions of disease- and care-related durations used in this OpenCOVID application of Switzerland.

**Figure S9:**
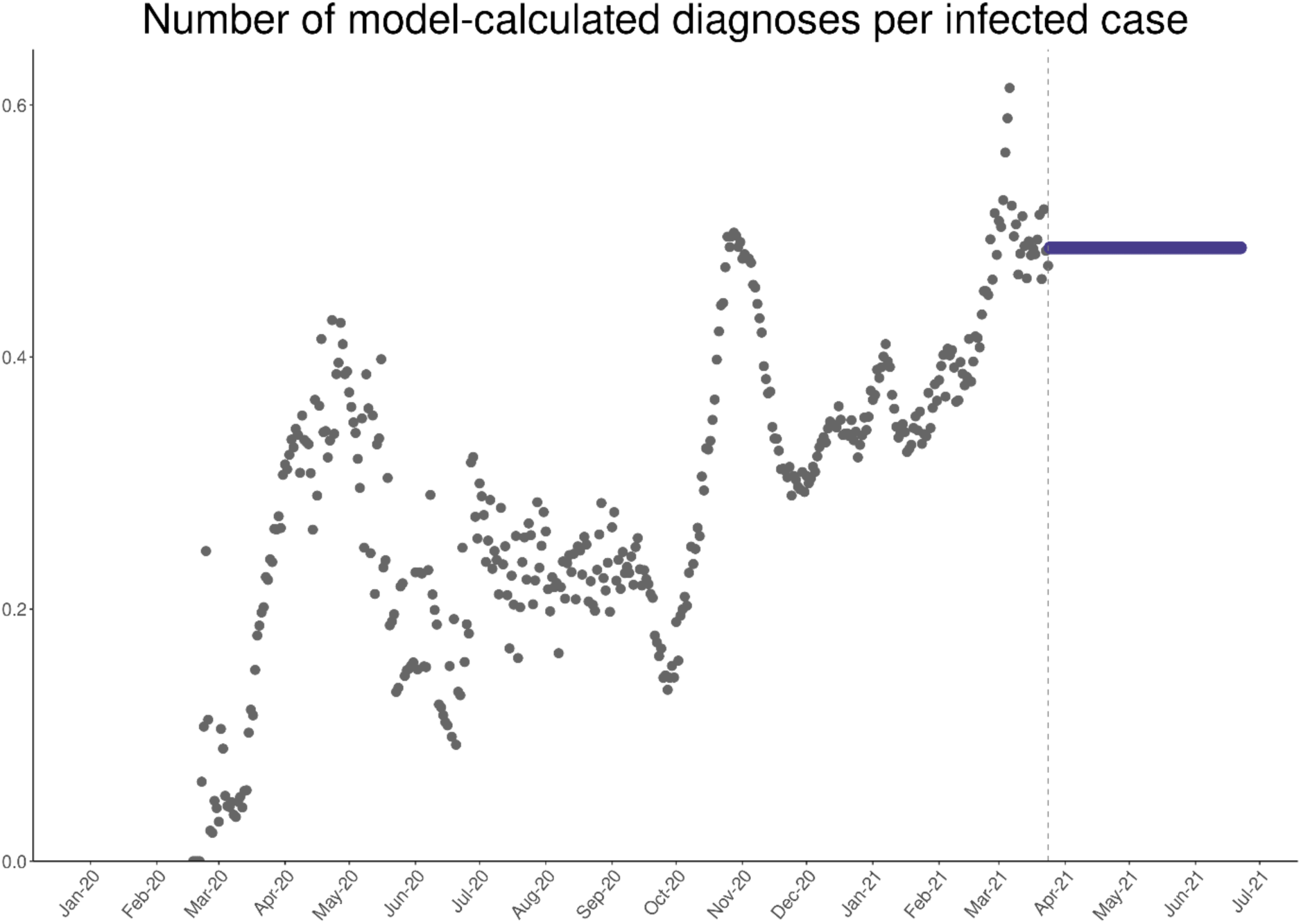
Proportion of modelled new infections to be diagnosed over time. The blue points represent the future testing and diagnosis assumption.

**Figure S10:**
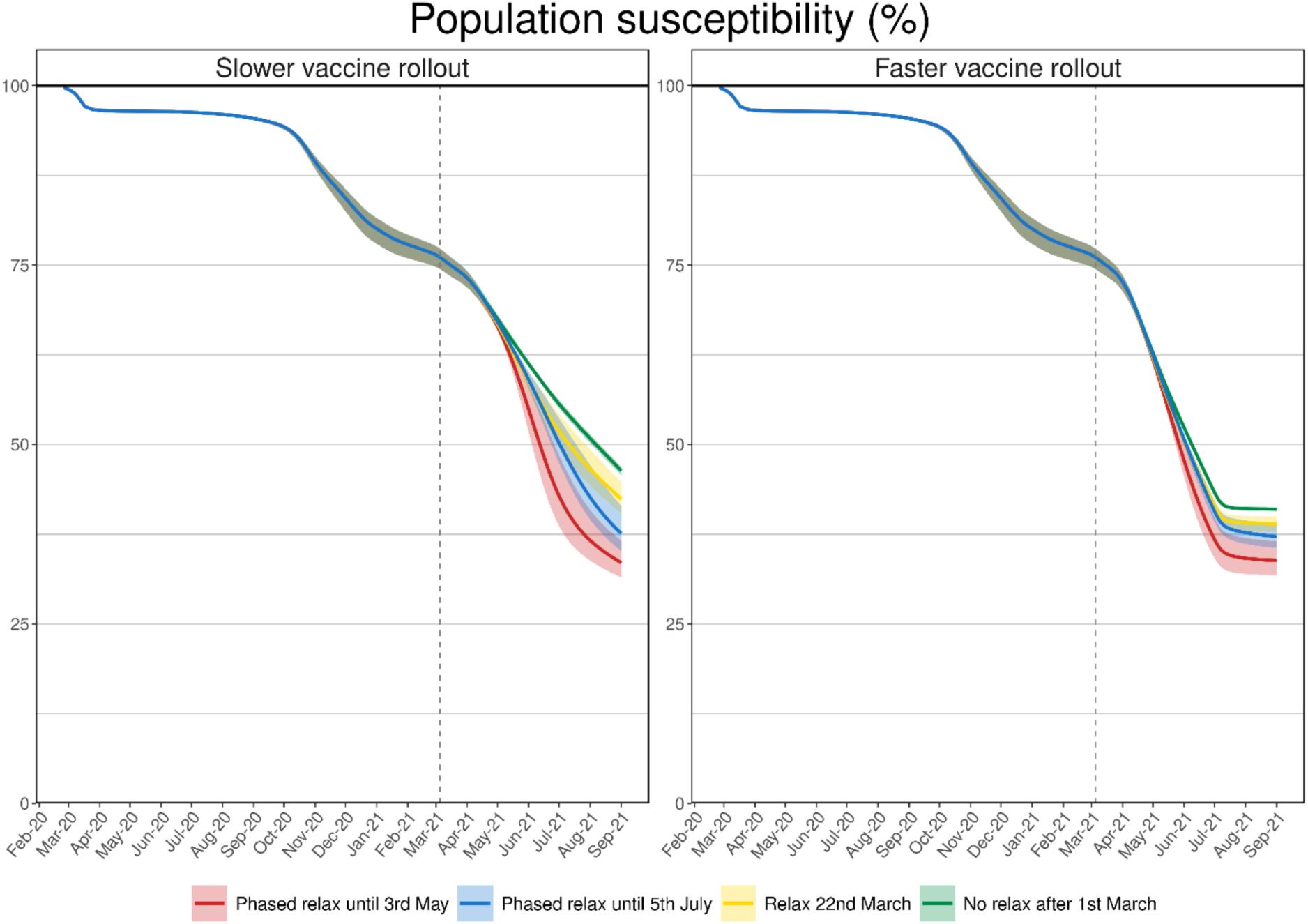
Development of population susceptibility levels of scenarios 1A-1D over time. Population susceptibility is the complement of population immunity. The colours are the scenarios from manuscript figure 2.

**Figure S11:**
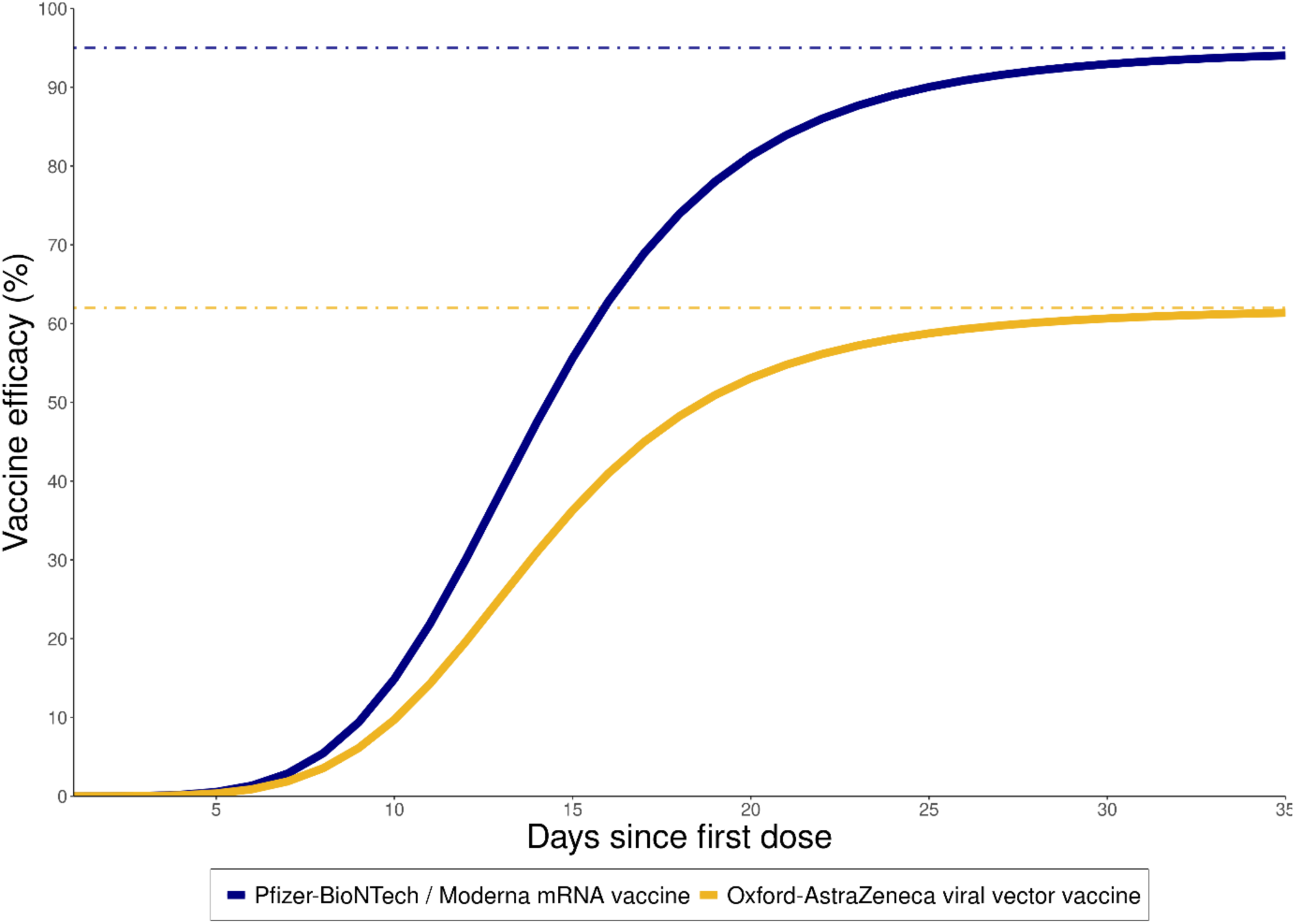
Development of vaccine efficacy from day of receiving a first dose. Shown in blue is the development of immunity for a two-dose course with an mRNA vaccine, and in yellow for a two-dose course of the AstraZeneca viral vector vaccine. The vaccine efficacy is the combined effect of immunity and the reduction of severe disease. It is assumed that the second dose is given according to schedule within 28 days depending on vaccine, but not modelled explicitly.

**Table S1:** Calibrated and fixed model parameters with sources.

| Model parameter | Description | Calibrated value or fixed distribution | Truncation bounds | Specifications and sources |
| --- | --- | --- | --- | --- |
| contacts | Number of contacts per person per day, where a contact has a transmission probability of beta | 11.4 | 5 – 25 | Calibrated<br>See *Contact network* section |
| beta | Baseline probability of transmission in one contact between a fully susceptible individual (i.e., no partial or sterile immunity) and a fully infectious individual (i.e., when viral load is at maximum) | $X = 0.05$ | Not applicable | <u>Not</u> calibrated<br>See *Infectiousness per contact* section |
| seasonality_scaler | Multiplicative factor for effect of temperature on transmission probability per contact | 0.27 | 0.2 – 0.8 | Calibrated<br>See *Seasonality* section |
| proportion_asymptomatic | Proportion of all cases that are asymptomatic | $X \sim N(0.3, 0.02^2)$ | 0.2 – 0.4 | <u>Not</u> calibrated<br>[4-9] |
| presymptomatic_days | Number of days infectious before showing symptoms | $X \sim N(3, 0.5^2)$ | 1 – 5 | <u>Not</u> calibrated<br>[10-14] |
| latency_days | Number of days in latency (infected but not infectious) state | $X \sim N(4.6, 1^2)$ | 3 – 7 | <u>Not</u> calibrated [10-14] |
| infectious_days_mild | Number of days for which non-severe cases are infectious (excluding presymptomatic phase) | $X \sim N(6, 1^2)$ | 3 – 10 | <u>Not</u> calibrated [10, 15-17] |
| infectious_days_severe | Number of days for which severe cases are infectious (excluding presymptomatic phase) | $X \sim N(28, 1^2)$ | 14 – 30 | <u>Not</u> calibrated [14, 15, 18-20] |
| seek_hospital | Proportion of severe cases that will seek hospital care | 0.41 | 0.4 – 0.95 | Calibrated<br>Assumed prior distribution [21] |
| onset_to_hospital_days | Number of days between symptom onset and hospitalisation | 13.4 | 1 – 14 | Calibrated [14, 20, 22-25] |
| diagnosis_delay | Delay between symptom onset and test (and diagnosis) | $X \sim N(3, 0.5^2)$ | 0 – 5 | <u>Not</u> calibrated [26] |
| reporting_delay | General reporting delay for all epidemiological and hospital metrics | $X \sim N(1, 0.1^2)$ | 0 – 4 | <u>Not</u> calibrated<br>Assumed |
| isolation_probability | Proportion of mild and asymptomatic cases that isolate after diagnosis | $X = 1$ | Not applicable | <u>Not</u> calibrated<br>Assumed |
| isolation_duration | Number of days spent in isolation after diagnosis | $X = 10$ | Not applicable | <u>Not</u> calibrated [17] |
| hospital_stay_days | Number of days a severe non-critical case spends in hospital before discharge | $X \sim N(9, 1^2)$ | 7 – 20 | <u>Not</u> calibrated [14, 20, 23, 25, 27] |
| hospital_to_icu_days | Number of days between hospital admission and ICU admission for cases that will become critical | $X \sim N(2, 1^2)$ | 1 – 10 | <u>Not</u> calibrated [23, 28] |
| icu_stay_days | Number of days a critical case spends in ICU before transfer back to non-ICU ward | $X \sim N(7, 1^2)$ | 1 – 14 | <u>Not</u> calibrated [14, 20, 23, 25, 27] |
| icu_stay_death_days | Number of days a critical case spends in ICU before death | $X \sim N(6, 1^2)$ | 3 – 14 | <u>Not</u> calibrated [14, 19, 20, 23, 27] |
| hospital_transfer_days | Number of days spent in non-ICU ward following discharge/transfer from ICU | $X \sim N(2, 0.1^2)$ | 1 – 7 | <u>Not</u> calibrated [22] |
| home_death_days | Number of days between symptom onset and death for those not seeking hospital care | $X \sim N(10, 1^2)$ | 7 – 14 | <u>Not</u> calibrated<br>Assumed |
| death_reporting_delay | Additional delay between a COVID-19 death and that death being reported in the data | $X \sim N(2, 0.2^2)$ | 1 – 14 | <u>Not</u> calibrated [22, 25] |
| critical_death_icu | Proportion of critical cases that die in ICU care (ventilators assumed to be available) | 0.63 | 0.4 – 0.8 | Calibrated [22, 24, 25] |
| critical_death_non_icu | Proportion of critical cases that die when ICU not available or not sought | $X \sim N(0.95, 0.1^2)$ | 0.5 – 0.99 | <u>Not</u> calibrated [25] |
| improved_care_factor | Proportionate reduction in probability of severe cases becoming critical cases (and requiring ICU) due to improved care and treatment procedures | 0.72 | 0.40 – 0.99 | Calibrated [29] |
| import_date | Number of days delay between first case importation and first confirmed case | $X = 7$ | Not applicable | <u>Not</u> calibrated<br>Correlated with import_initial |
| import_initial | Number of people (per 100,000) initiated with infection import_date number of days before first confirmed cases | 78 | 1 – 100 | Calibrated<br>Assumed prior distribution |
| import_constant | Number of imported cases per 100,000 people per day | 1.4 | 0.1 – 5 | Calibrated<br>Assumed prior distribution |
| npi_scaler | Calibration factor for proportionally scaling the Oxford Health and Containment Index (OCHI) to derive a reduction in effective contacts | 1.3 | 0.8 – 1.5 | Calibrated<br>See *Non-pharmaceutical interventions* section |
| acquired_immunity | Initial level of acquired immunity due to infection | $X \sim N(0.83, 0.1^2)$ | 0.76 – 0.87 | <u>Not</u> calibrated<br>[30, 31] |
| vaccine_immunity_days | Number of days following vaccination (first dose) until full efficacy is attained | $X = 28$ | No bounds assumed | <u>Not</u> calibrated<br>[32] |

**Table S2:** Baseline properties of viral variants modelled for this application of Switzerland. * Note that the sensitivity of model outputs to varying infectivity and disease severity factors were quantified in a sensitivity analysis.

| Viral variant | Model import date | Number imported (per 100,000 people) | Infectivity factor | Disease severity factor |
| --- | --- | --- | --- | --- |
| D614 | See Table S1 | Calibrated (see Table S1) | 1<br>*Reference* | 1<br>*Reference* |
| B.1.1.7 | 24 November 2020 | 12 | 1.6* [1.5-1.7] | 1* [up to 1.5] |
| B.1.351 | 1 December 2020 | 8 | 1.1 | 1 |

**Table S3:** Weightings of calibration metrics in likelihood function

| Metric | Weighting ($\omega_i$) | Comments |
| --- | --- | --- |
| Daily confirmed cases | 1 | Default weighting |
| Daily COVID-19 deaths | 2 | Higher weighting to account for higher reporting probability |
| New daily admissions to hospital | 0 | No weighting applied due to discrepancies between data sources |
| COVID-19 cases in hospital | 4 | Higher weighting to account for higher confidence in reporting |
| COVID-19 cases in ICU | 4 | Higher weighting to account for higher confidence in reporting |
| Variant prevalence | 1.6 | Weighting of 50 applied to 12 data points implies a relative weighting of 1.6 |

**Table S4:** Age-group dependant probabilities of a given prognosis.

| Age group | Asymptomatic | Mild disease | Severe disease | Critical disease | Death |
| --- | --- | --- | --- | --- | --- |
| **0-10** | 30% | 69.93% | 0.07% | 0.00% | <0.0001% |
| **10-20** | 30% | 69.79% | 0.20% | 0.01% | <0.0001% |
| **20-30** | 30% | 69.16% | 0.80% | 0.04% | <0.0001% |
| **30-40** | 30% | 67.76% | 2.13% | 0.11% | 0.0002% |
| **40-50** | 30% | 66.57% | 3.21% | 0.21% | 0.0014% |
| **50-60** | 30% | 62.86% | 6.27% | 0.85% | 0.0193% |
| **60-70** | 30% | 58.38% | 8.44% | 2.95% | 0.23% |
| **70-80** | 30% | 52.99% | 9.66% | 5.56% | 1.79% |
| **80-90+** | 30% | 50.89% | 5.56% | 1.63% | 11.92% |

**Table S5:** Vaccine properties under different assumptions of vaccine transmission blocking effect.

| Vaccine type | Overall efficacy ($\theta$) | Sterile immunity effect ($\alpha$) | Symptom reducing effect ($\gamma$) |
| --- | --- | --- | --- |
| mRNA | 95% | 80% | 75% |
| mRNA | 95% | 60% | 87.5% |
| mRNA | 95% | 95% | 0% |

**Table S6:** Vaccine priority groups based on FOPH priority groups and modified after discussion with FOPH.

| Priority group | Description | Number of model-eligible people |
| --- | --- | --- |
| **P1** | Aged over 75 years | 756,400 |
| **P2** | Aged between 65 and 75 years, under 65 with comorbidities, and healthcare workers | 2,031,600<br>(850,000, 621,600, and 560,000, respectively) |
| **P3** | Household members of high-risk people | 1,243,000 |
| **P4** | Adults (18-64 years) in communal facilities and their caregivers | 100,000 |
| **P5** | All other adults (18-65 years) | 4,414,600 |

## Notes

### Competing Interest Statement

The authors have declared no competing interest.

## References

1. BAG. Coronavirus-Krankheit-2019 (Covid-19) – Stand: 01.02.2021, 08.04h Situationsbericht zur epidemiologischen Lage in der Schweiz und im Fürstentum Liechtenstein, https://www.covid19.admin.ch (2021).

2. Easing and tightening of nationwide measures, https://www.bag.admin.ch/dam/bag/en/dokumente/mt/k-und-i/aktuelle-ausbrueche-pandemien/2019-nCoV/covid-19-tabelle-lockerung.pdf.download.pdf/Easing_of_measures_and_possible_next_steps.pdf (2020).

3. Flaxman, S. et al. Estimating the effects of non-pharmaceutical interventions on COVID-19 in Europe. Nature, Preprint at https://doi.org/10.1038/s41586-020-2405-7 (2020).

4. Scott, N. et al. Modelling the impact of reducing control measures on the COVID-19 pandemic in a low transmission setting. Med. J. Aust. (2020).

5. Moore, S., Hill, E. M., Tildesley, M. J., Dyson, L. & Keeling, M. J. Vaccination and non-pharmaceutical interventions for COVID-19: a mathematical modelling study. Lancet Infect. Dis. (2021).

6. Hale, T. et al. A global panel database of pandemic policies (Oxford COVID-19 Government Response Tracker). Nature Human Behaviour, 1–10 (2021).

7. Stringhini, S. et al. Seroprevalence of anti-SARS-CoV-2 antibodies after the second pandemic peak. Lancet Infect. Dis., Preprint at https://doi.org/10.1016/S1473-3099(21)00054-2 (2021).

8. West, E. A. et al. Corona Immunitas: study protocol of a nationwide program of SARS-CoV-2 seroprevalence and seroepidemiologic studies in Switzerland. Int. J. Public Health 65, 1529–1548, Preprint at https://doi.org/10.1007/s00038-020-01494-0 (2020).

9. Binois, M., Gramacy, R. B. & Ludkovski, M. Practical heteroscedastic gaussian process modeling for large simulation experiments. J. Comp. Graph. Stat. 27, 808–821 (2018).

10. Chen, C. et al. Quantification of the spread of SARS-CoV-2 variant B.1.1.7 in Switzerland. Preprint at https://doi.org/10.1101/2021.03.05.21252520 (2021).

11. Davies, N. G. et al. Estimated transmissibility and impact of SARS-CoV-2 lineage B. 1.1. 7 in England. Science (2021).

12. Reichmuth, M. et al. Transmission of SARS-CoV-2 variants in Switzerland, https://ispmbern.github.io/covid-19/variants/ (2021).

13. Quantification-of-the-spread-of-a-SARS-CoV-2-variant, https://github.com/cevo-public/Quantification-of-the-spread-of-a-SARS-CoV-2-variant (2021).

14. COVID-19 Switzerland: Hospital capacity, ICUs, https://www.covid19.admin.ch/en/hosp-capacity/icu (2021).

15. Davies, N. G. et al. Increased mortality in community-tested cases of SARS-CoV-2 lineage B.1.1.7. Nature, Preprint at https://doi.org/10.1038/s41586-021-03426-1 (2021).

16. Challen, R. et al. Risk of mortality in patients infected with SARS-CoV-2 variant of concern 202012/1: matched cohort study. BMJ 372, n579, Preprint at https://doi.org/10.1136/bmj.n579 (2021).

17. Cheng, C. et al. icumonitoring.ch: a platform for short-term forecasting of intensive care unit occupancy during the COVID-19 epidemic in Switzerland. Swiss Med. Wkly. 150, w20277 (2020).

18. National COVID-19 Science Task Force Effect of the measures 17th April 2020. (https://sciencetaskforce.ch/policy-brief/effect-of-measures/, 2020).

19. Christie, A., Mbaeyi, S. A. & Walensky, R. P. CDC Interim Recommendations for Fully Vaccinated People: An Important First Step. JAMA (2021).

20. Fontanet, A. et al. SARS-CoV-2 variants and ending the COVID-19 pandemic. Lancet (2021).

21. Madhi, S. A. et al. Efficacy of the ChAdOx1 nCoV-19 Covid-19 vaccine against the B.1.351 variant. N. Engl. J. Med. (2021).

22. Byrne, A. W. et al. Inferred duration of infectious period of SARS-CoV-2: rapid scoping review and analysis of available evidence for asymptomatic and symptomatic COVID-19 cases. BMJ Open 10, e039856 (2020).

23. Prevalence of long COVID symptoms and COVID-19 complications, https://www.ons.gov.uk/peoplepopulationandcommunity/healthandsocialcare/healthandlifeexpectancies/datasets/prevalenceoflongcovidsymptomsandcovid19complications (2020).

24. Carfì, A., Bernabei, R. & Landi, F. Persistent symptoms in patients after acute COVID-19. JAMA 324, 603–605 (2020).

25. COVID measures by canton, Swiss Tropical and Public Health Institute, https://github.com/SwissTPH/COVID_measures_by_canton (2021).

26. Hall, V. J. et al. Do antibody positive healthcare workers have lower SARS-CoV-2 infection rates than antibody negative healthcare workers? Large multi-centre prospective cohort study (the SIREN study), England: June to November 2020. medRxiv, 2021.2001. 2013.21249642 (2020).

27. Polack, F. P. et al. Safety and efficacy of the BNT162b2 mRNA Covid-19 vaccine. N. Engl. J. Med. 383, 2603–2615 (2020).

28. Baden, L. R. et al. Efficacy and safety of the mRNA-1273 SARS-CoV-2 vaccine. N. Engl. J. Med. 384, 403–416 (2021).

29. Mossong, J. et al. Social contacts and mixing patterns relevant to the spread of infectious diseases. PLoS Med. 5, e74 (2008).

30. BAG. https://www.bag.admin.ch/dam/bag/de/dokumente/mt/k-und-i/aktuelle-ausbrueche-pandemien/2019-nCoV/Unterlagen-Konsultationen-Kantone/begleitdokument-bes-lage-lockerung-1.pdf.download.pdf/Begleitdokument%20f%C3%BCr%20die%20Kantone.pdf (2021).

31. CH Covid-19 Dashboard, https://ibz-shiny.ethz.ch/covidDashboard/ (2021).

32. COVID-19 Switzerland, Status report, Switzerland and Liechtenstein, https://www.covid19.admin.ch/en/overview (2021).

33. CH Meteo Schweiz, https://data.geo.admin.ch/ch.meteoschweiz.klima/normwerte/normwerte.zip (2021).

34. Thomas Hale SW, Petherick A, Phillips T & Kira B. https://www.bsg.ox.ac.uk/research/research-projects/coronavirus-government-response-tracker (University of Oxford, Blavatnik School of Government, Oxford, United Kingdom, 2020).

35. Moderna. A phase 3, randomized, stratified, observer-blind, placebo-controlled study to evaluate the efficacy, safety, and immunogenicity of mRNA-1273 SARS-CoV-2 vaccine in adults aged 18 years and older. https://clinicaltrials.gov/ct2/show/NCT04470427 (2020).

36. BAG. COVID-19 Switzerland, key figures, Switzerland and Liechtenstein, vaccine doses, https://www.covid19.admin.ch/en/epidemiologic/vacc-doses (2021).

37. BAG. Covid-19-Impfstrategie, https://www.bag.admin.ch/dam/bag/de/dokumente/mt/k-und-i/aktuelle-ausbrueche-pandemien/2019-nCoV/impfstrategie-bag-ekif.pdf.download.pdf/COVID-19_Impfstrategie_BAG-EKIF_Stand%2016.12.20.pdf (2020).

38. BAG. Weiteres Vorgehen bezüglich der nationalen Massnahmen, https://www.google.com/search?q=BAG+Weiteres+Vorgehen+bez%C3%BCglich+der+nationalen+Massnahme&rlz=1C1CHBF_enCA922CA922&oq=BAG+Weiteres+Vorgehen+bez%C3%BCglich+der+nationalen+Massnahme&aqs=chrome..69i57l2j69i59l2j69i60j69i61l2j69i60.1166j0j7&sourceid=chrome&ie=UTF-8 (2021).

## References

1. FOPH COVID-19 Switzerland Information on the current situation. Available from: https://www.covid19.admin.ch/en/overview.

2. Quantification-of-the-spread-of-a-SARS-CoV-2-variant. Available from: https://github.com/cevo-public/Quantification-of-the-spread-of-a-SARS-CoV-2-variant.

3. Chen, C., et al., Quantification of the spread of SARS-CoV-2 variant B.1.1.7 in Switzerland. medRxiv, 2021: p. 2021.03.05.21252520.

4. Nishiura, H., et al., Estimation of the asymptomatic ratio of novel coronavirus (2019- nCoV) infections among passengers on evacuation flights. medRxiv, 2020.

5. Mizumoto, K., et al., Estimating the asymptomatic proportion of coronavirus disease 2019 (COVID-19) cases on board the Diamond Princess cruise ship, Yokohama, Japan, 2020. Euro Surveill, 2020. 25(10): p. 2000180.

6. Qiu, J., Covert coronavirus infections could be seeding new outbreaks. 2020.

7. Wu, J., et al., Identification of RT-PCR-Negative Asymptomatic COVID-19 Patients via Serological Testing. Front Public Health, 2020. 8: p. 267.

8. Byambasuren, O., et al., Estimating the extent of true asymptomatic COVID-19 and its potential for community transmission: systematic review and meta-analysis. 2020.

9. Gudbjartsson, D.F., et al., Spread of SARS-CoV-2 in the Icelandic Population. N Engl J Med, 2020. 382(24): p. 2302–2315.

10. Kissler, S.M., et al., Projecting the transmission dynamics of SARS-CoV-2 through the postpandemic period. Science, 2020. 368(6493): p. 860–868.

11. Read, J.M., et al., Novel coronavirus 2019-nCoV: early estimation of epidemiological parameters and epidemic predictions. MedRxiv, 2020.

12. Lauer, S.A., et al., The Incubation Period of Coronavirus Disease 2019 (COVID-19) From Publicly Reported Confirmed Cases: Estimation and Application. Ann Intern Med, 2020. 172(9): p. 577–582.

13. Lei, S., et al., Clinical characteristics and outcomes of patients undergoing surgeries during the incubation period of COVID-19 infection. E Clinical Medicine, 2020: p. 100331.

14. Zhao, C., et al., Public health initiatives from hospitalized patients with COVID-19, China. J Infect Public Health, 2020.

15. He, X., et al., Temporal dynamics in viral shedding and transmissibility of COVID-19. Nat Med, 2020. 26(5): p. 672–675.

16. Du, Z., et al., Serial Interval of COVID-19 among Publicly Reported Confirmed Cases. Emerg Infect Dis, 2020. 26(6): p. 1341–1343.

17. Woelfel, R., et al., Clinical presentation and virological assessment of hospitalized cases of coronavirus disease 2019 in a travel-associated transmission cluster. MedRxiv, 2020.

18. Peirlinck, M., et al., Outbreak dynamics of COVID-19 in China and the United States. Biomech Model Mechanobiol, 2020: p. 1.

19. Zhao, W., et al., Clinical characteristics and durations of hospitalized patients with COVID-19 in Beijing: a retrospective cohort study. MedRxiv, 2020.

20. Hua, J., et al., Epidemiological features and medical care-seeking process of patients with COVID-19 in Wuhan, China. ERJ Open Res, 2020. 6(2).

21. Federal Office of Public Health FOPH. COVID-19 Switzerland: Status report, Switzerland and Liechtenstein (last updated 22 March 2021). [cited 2021 22 March]; Available from: https://www.covid19.admin.ch/en/overview.

22. European Centre for Disease Prevention and Control, The European Surveillance System (TESSy). 2020: Stockholm.

23. Linton, N.M., et al., Incubation Period and Other Epidemiological Characteristics of 2019 Novel Coronavirus Infections with Right Truncation: A Statistical Analysis of Publicly Available Case Data. J Clin Med, 2020. 9(2): p. 538.

24. Federal Office of Public Health. Federal Council of Switzerland, Neues Coronavirus: Situation Schweiz. 2020: Bern.

25. European Centre for Disease Prevention and Control, Projected baselines of COVID- 19 in the EU/EEA and the UK for assessing the impact of de-escalation of measures. 2020, ECDC: Stockholm.

26. Kretzschmar, M.E., et al., Impact of delays on effectiveness of contact tracing strategies for COVID-19: a modelling study. Lancet Public Health, 2020. 5: p. e452–e459.

27. Verity, R., et al., Estimates of the severity of coronavirus disease 2019: a model-based analysis. Lancet Infect Dis, 2020. 20(6): p. 669–677.

28. Thai, P.Q., et al., Factors associated with the duration of hospitalisation among COVID-19 patients in Vietnam: A survival analysis. Epidemiol Infect, 2020. 148: p. e114.

29. The RECOVERY Collaborative Group, Dexamethasone in Hospitalized Patients with Covid-19. New England Journal of Medicine, 2020. 384(8): p. 693–704.

30. Hall, V., et al., Do antibody positive healthcare workers have lower SARS-CoV-2 infection rates than antibody negative healthcare workers? Large multi-centre prospective cohort study (the SIREN study), England: June to November 2020. medRxiv, 2021: p. 2021.01.13.21249642.

31. Mahase, E., Covid-19: Past infection provides 83% protection for five months but may not stop transmission, study finds. BMJ, 2021. 372: p. n124.

32. Hunter, P.R. and J. Brainard, Estimating the effectiveness of the Pfizer COVID-19 BNT162b2 vaccine after a single dose. A reanalysis of a study of ‘real-world’ vaccination outcomes from Israel. medRxiv, 2021: p. 2021.02.01.21250957.

33. sciCORE scientific computing core facility at University of Basel. Available from: https://scicore.unibas.ch/.

34. Binois, M., R.B. Gramacy, and M. Ludkovski, Practical heteroscedastic gaussian process modeling for large simulation experiments. Journal of Computational and Graphical Statistics, 2018. 27(4): p. 808–821.

35. Binois, M. and R. Gramacy, hetGP: Heteroskedastic Gaussian process modeling and sequential design in R. 2019.

36. Mossong, J., et al., POLYMOD social contact data. 2017.

37. Funk, S., Socialmixr: Social mixing matrices for infectious disease modelling. 2018.

38. Ferretti, L., et al., The timing of COVID-19 transmission. 2020.

39. Kissler, S.M., et al., Viral dynamics of SARS-CoV-2 infection and the predictive value of repeat testing. medRxiv, 2020.

40. Climatology, F.O.o.M.a. MeteoSwiss. 2021; Available from: https://www.meteoswiss.admin.ch/home.html?tab=overview.

41. Office, F.S. Swiss Open Government data. 2021; Available from: https://opendata.swiss/en.

42. Office, F.S. Climatological Network - Daily Values. 2021; Available from: https://opendata.swiss/en/dataset/klimamessnetz-tageswerte.

43. Cordano, E., The ‘RMAWGEN’ package for the R programming language.

44. Hale, T., et al., Variation in government responses to COVID-19. Blavatnik school of government working paper, 2020. 31: p. 2020–11.

45. Hale, T. and S. Webster, Oxford COVID-19 government response tracker. 2020.

46. COVID measures by canton. 2021; Available from: https://github.com/SwissTPH/COVID_measures_by_canton.

47. BAG. Available from: https://www.bag.admin.ch/dam/bag/de/dokumente/mt/k-und-i/aktuelle-ausbrueche-pandemien/2019-nCoV/Unterlagen-Konsultationen-Kantone/begleitdokument-bes-lage-lockerung-1.pdf.download.pdf/Begleitdokument%20f%C3%BCr%20die%20Kantone.pdf.

48. Available from: https://ibz-shiny.ethz.ch/covidDashboard/.

49. Good, M.F. and M.T. Hawkes, The Interaction of Natural and Vaccine-Induced Immunity with Social Distancing Predicts the Evolution of the COVID-19 Pandemic. Mbio, 2020. 11(5).

50. Spellberg, B., T.B. Nielsen, and A. Casadevall, Antibodies, immunity, and COVID-19. JAMA internal medicine, 2020.

51. Edridge, A.W., et al., Seasonal coronavirus protective immunity is short-lasting. Nature medicine, 2020. 26(11): p. 1691–1693.

52. Ihaka, R. and R. Gentleman, R: a language for data analysis and graphics. Journal of computational and graphical statistics, 1996. 5(3): p. 299–314.

53. OpenCOVID soure code. 2021; Available from: https://github.com/SwissTPH/OpenCOVID.

